# Non-stationary Spatio-Temporal Modeling of COVID-19 Progression in The U.S.

**DOI:** 10.1101/2020.09.14.20194548

**Authors:** Yue Bai, Abolfazl Safikhani, George Michailidis

## Abstract

The fast transmission rate of COVID-19 worldwide has made this virus the most important challenge of year 2020. Many mitigation policies have been imposed by the governments at different regional levels (country, state, county, and city) to stop the spread of this virus. Quantifying the effect of such mitigation strategies on the transmission and recovery rates, and predicting the rate of new daily cases are two crucial tasks. In this paper, we propose a modeling framework which not only accounts for such policies but also utilizes the spatial and temporal information to characterize the pattern of COVID-19 progression. Specifically, a piecewise susceptible-infected-recovered (SIR) model is developed while the dates at which the transmission/recover rates change significantly are defined as “break points” in this model. A novel and data-driven algorithm is designed to locate the break points using ideas from fused lasso and thresholding. In order to enhance the forecasting power and to describe additional temporal dependence among the daily number of cases, this model is further coupled with spatial smoothing covariates and vector auto-regressive (VAR) model. The proposed model is applied to several U.S. states and counties, and the results confirm the effect of “stay-at-home orders” and some states’ early “re-openings” by detecting break points close to such events. Further, the model performed satisfactorily short-term forecasts of the number of new daily cases at regional levels by utilizing the estimated spatio-temporal covariance structures. Finally, some theoretical results and empirical performance of the proposed methodology on synthetic data are reported which justify the good performance of the proposed method.

## 1. Introduction

Since the first officially reported case in China in late December 2019, the SARS-CoV-2 virus (henceforth COVID-19) spread worldwide within weeks. As of August 18, 2020, there have been over five million confirmed cases of COVID-19 in the United States alone and more than twenty million worldwide. In response to the rapid growth of confirmed cases, followed by hospitalizations and fatalities, initially in the Hubei Province and in particular the capital of Wuhan in China, and subsequently in Northern Italy, Spain and the tri-state area of New York, New Jersey and Connecticut, various mitigation strategies were put rapidly in place with the most stringent one of “stay-at-home” orders. The key purpose of such strategies was to reduce the virus transmission rate and reduce pressure on public health infrastructure (Anderson et al., 2020). To that end, the California governor issued a “stay-at-home” order on March 19, 2020, that was quickly followed by another 42 states by early April. All states with such orders proceeded with multi-phase reopening plans starting in early May, allowing various non-essential business to operate, possibly at reduced capacity levels to enforce social distancing guidelines. In addition, mask wearing mandates also came into effect (CDC, 2020) as emerging evidence from clinical and laboratory studies shows that masks reduce the spread (Leung et al., 2020). However, these reopening plans led to a substantial increase in the number of confirmed COVID-19 cases in many US states, followed by increased number of fatalities.

The emergence of the COVID-19 worldwide pandemic led to the development of many modeling approaches for both prediction purposes and also assessing various mitigation strategies. Next, a brief overview of proposed models is provided.

*Related Work*. A number of epidemic models have been developed to analyze and predict COVID-19 transmission dynamics. Mathematical models, such as the class of susceptible-infectious-recovered (SIR) models are widely used to model and forecast epidemic spreads. Chen, Lu and Chang (2020) proposed a time-dependent SIR model and tracked the transmission rate and the recovery rate at each time point by employing ridge regression while Anastassopoulou et al. (2020) proposed a discrete-time susceptible-infectious-recovered-dead (SIRD) model and provided estimations of the basic reproduction number (Ro), and the infection mortality and recovery rates by least squares method. Moreover, Song et al. (2020) built an extended SIR model with time-varying transmission rates and implemented the MCMC algorithm to obtain posterior estimates and credible intervals of the unknown parameters

A number of models focused on identifying a change in the parameters of the underlying model employed. For example, Dehning et al. (2020) combined the widely used SIR model (see Section 2) with Bayesian parameter inference through Markov Chain Monte Carlo (MCMC), assuming a time-dependent transmission rate. Instead of directly estimating a change point in the transmission rate and the other parameters in the SIR model, they assumed a fixed value on the number of the change points, and imposed informative prior distributions on their locations, as well as the transmission rate based on information from intervention policies. Further, Jiang, Zhao and Shao (2020) proposed to model the time series of the log-scaled cumulative confirmed cases and deaths of each country via a piecewise linear trend model. They combined the self-normalization (SN) change-point test with the narrowest-over-threshold (NOT) algorithm (Baranowski, Chen and Fryzlewicz, 2016) to achieve multiple change-point estimation. Moreover, Vokó and Pitter (2020) and Wagner et al. (2020) analyzed the effect of social distancing measures adopted in Europe and the United States, respectively, using an interrupted time series (ITS) analysis of the confirmed case counts. Their work aim to find a change point in the time series data of confirmed cases counts for which there is a significant change in the growth rate. In Vokó and Pitter (2020)’s paper, the change points were determined by linear threshold regression models of the logarithm of daily cases while Wagner et al. (2020) used an algorithm developed in Fearnhead, Maidstone and Letchford (2019), based on an *L*_0_ penalty on changes in slope to identify the change points.

Another line of work employed spatio-temporal models for parameter estimation and forecasting the spread of COVID-19. For example, Wang et al. (2020a) introduced a generalized additive varying coefficient model and coupled it with a non-parametric approach for modeling the data, to study spatio-temporal patterns in the spread of COVID-19 at the county level. Further, Srivastava and Prasanna (2020) proposed a heterogeneous infection rate model with human mobility from multiple regions and trained it using weighted least squares at state and country levels while Qi et al. (2020) fitted a Generalized Additive Model (GAM) to quantify the province-specific associations between meteorological variables and the daily cases of COVID-19 during the period under consideration.

In addition to mathematical methods, many machine learning/deep learning methods were applied for forecasting of COVID-19 transmission. For example, Chimmula and Zhang (2020) employed Long Short-Term Memory (LSTM) type deep neural networks to forecast the future COVID-19 cases in Canada while Hu et al. (2020) developed a modified stacked auto-encoder for modeling the transmission dynamics of the epidemic.

In this paper, we analyze the confirmed and death cases data related to COVID-19 from five states and nine parishes/counties/cities in the United States from March 1st to August 18, 2020. In the absence of interventions, the spread of COVID-19 can be modeled by a Susceptible-Infected-Recovered (SIR) model with stationary transmission and recovery rates. One of the main reasons to choose the SIR model as the building block for our proposed model is that the transmission and recovery rates in the SIR model are easy to interpret and hence can be utilized in policy decision making process. However, since there were many mitigation policies put in place at different regional levels in the U.S., the simple SIR model may not be a good fit. Instead, we propose a piecewise stationary SIR model (model 1), i.e. the SIR model parameters may change at certain (unknown) time points. Such time points are defined as “break (change) points”. Unlike some other methods discussed in the literature review, in our modeling framework, the number of change points and their locations are assumed to be *unknown* and must be inferred from the data. Such flexibility in the modeling end not only allows inferring potentially different temporal patterns in different regions (states or counties) but also complicates the model fitting procedures. To that end, a novel data-driven algorithm is developed to detect all break points, and to estimate the model parameters within each stationary segment. Specifically, we define certain time blocks and assume the model parameters are fixed during each block of time points. Then, fused lasso based estimation is utilized to estimate all model parameters (Tibshirani et al., 2005). This procedure is further coupled with hard-thresholding and exhaustive search steps to estimate the number and location of change points (see more details in Section 2, Algorithm 1). To enhance the forecasting power of the model and to capture additional spatial and temporal dependence not explained through the SIR model, the piecewise constant SIR model is coupled with spatial smoothing (model 2) and time series components (model 3). The former is done through adding a spatial effect term which accounts for the effect of neighboring regions while the latter includes adding a Vector Auto-Regressive (VAR) model component to capture additional auto-correlations among new daily new cases and deaths. Capturing the spatio-temporal dependence through model 3 helped in reducing the prediction error significantly (sometimes around 80%) compared to the piecewise SIR model which confirms the usefulness of such modeling framework (see more details in Section 4). To verify the applicability of the proposed methodology to other data sets with similar characteristics, the developed algorithm is tested over several simulation settings with satisfactory performance (see details in Section 3) and some theoretical properties of the proposed method (prediction consistency as well as detection accuracy) are investigated and summarized in Appendix A.

The rest of the paper is organized as follows. In Section 2, three statistical models are introduced and data-driven algorithms are described to estimate their parameters. The proposed algorithms are tested on various simulation settings and the results are reported in Section 3. The proposed models are applied to several U.S. states and counties and the results are described in Section 4. Finally, some concluding remarks are drawn in Section 5.

## 2. A Family of Spatio-temporal Heterogeneous SIR models

The proposed class of models leverages the framework of the SIR model, which is presented next to set up key concepts.

*Background: the standard SIR Model with Stationary Transmission and Recovery Rates:* We start by considering the standard SIR model (Kermack and McKendrick, 1927), wherein the total population is divided into the following three compartments: susceptible (uninfected), infected, and recovered (healed or dead). It is assumed that each infected individual, on average, infects *β* other individuals per unit time, and each infected individual recovers at rate *γ*. The two key model parameters, the transmission rate *β* and recovery rate *γ*, are assumed to be fixed over time. The temporal evolution of the SIR model is governed by the following system of three ordinary differential equations:

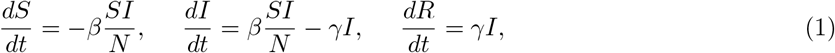

where *S*, *I* and *R* represent the individuals in the population in the susceptible, infected and recovered stages, respectively. Note that the variables *S*, *I* and *R* always satisfy *S* + *I* + *R* = *N*, where *N* is the total population size. In this formulation, we ignore the change in the total population, so that *N* remains constant over time. We also omit the probability rate of becoming susceptible again, after having recovered from the infection. Due to the fact that COVID-19 records are discrete in time (*Δ_t_* = 1 day), we consider the discrete-time version of SIR model, so that for each *t* =1,…,*T* − 1, the system comprises of the following three difference equations

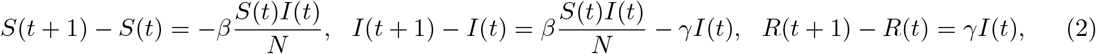

where *S*(*t*) stand for the number of susceptible individuals at time *t*, *I*(*t*) for the number of infected ones and finally *R*(*t*) for those recovered. Note that these three variables *S*(*t*), *I*(*t*) and *R*(*t*) still satisfy the constraint *S* + *I* + *R* = *N*. In the discrete version of the model, it is assumed that the observation period *T* is relatively small, so that only a very small fraction of the population is infected or recovered, i.e. *S*(*t*) ≫ *I*(*t*) and *S*(*t*) ≫ *R*(*t*). Therefore, we have *S*(*t*) ≈ *N* for all *t* ∈ [1,*T*]. Due to the constraint *S* + *I* + *R* = *N* and the assumption that *S*(*t*)*/N* ≈ 1, we drop variable *S*(*t*) and only consider the remaining two variables *I*(*t*) and *R*(*t*). In that case, for each *t* =1,…,*T* − 1, the difference equations reduce to the following simple linear equations:

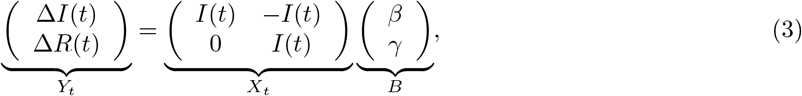

where *ΔI*(*t*)= *I*(*t* + 1) − *I*(*t*) and *ΔR*(*t*)= *R*(*t* + 1) − *R*(*t*).

Next, we extend the standard SIR model to accommodate temporal and spatial heterogeneity. The former is achieved, by allowing the transmission and recovery rates to vary over time, while the latter through the inclusion of an additional term in (3) that captures spatial effects.

### 2.1. Model 1: Piecewise Stationary SIR Model

The assumptions underlying the transmission and recovery rates of the standard SIR model are stringent. Both environmental factors and changes in population behavior can lead to time varying behavior and this has been the case for Covid-19; see, e.g., discussion in (Giordano et al., 2020). Variants of the SIR model with time varying parameters have been proposed in the literature (Liu and Stechlinski, 2012).

Let *β*(*t*) and *γ*(*t*) denote the transmission and recovery rate at time *t*. A time varying analogue of the system in (3) is:

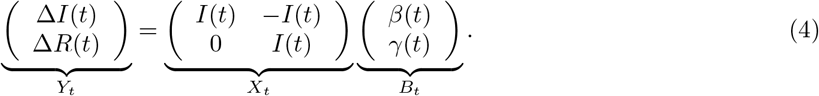

The solutions for *β*(*t*) and γ(*t*) are given by

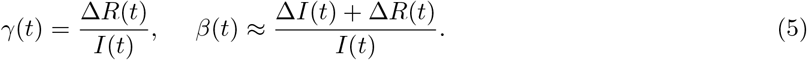

Given time series data on {*I*(*t*),*R*(*t*)),*t* ∈ [1,*T*]}, the corresponding *B_t_* =(*β*(*t*),*γ*(*t*))‣ can be calculated for each *t* =1, ···, *T* − 1 (Chen, Lu and Chang, 2020).

For our application, we assume that the transmission and recovery rates are *piecewise constant* over time, reflecting the fact that their temporal evolution is impacted by intervention strategies and environmental factors. Hence, it becomes important to identify the *break points* wherein these rates exhibit changes. Specifically, suppose there exist *m*_0_ change points 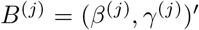 such that the rate vector *B_t_* =(*β*(*t*),*γ*(*t*))′ exhibits a change from *B*^(^*^j^*^)^=(*β*^(^*^j^*^)^,*γ*^(^*^j^*^)^)′ to *B*^(^*^j^*^+1)^=(*β*^(^*^j^*^+1)^,*γ*^(^*^j^*^+1)^)‣ at time point *t_j_*, while it remains fixed until the next break point. Then, model (4) takes the following form

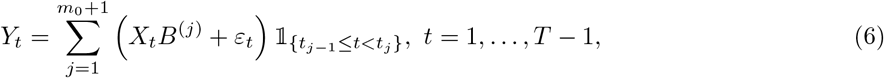

where 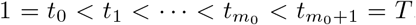. In each segment [*t_j_*_−1_,*t_j_*), all model parameters are assumed to be fixed; hence, these break points divide the time series data into stationary segments, as reflected in model (6).

The goal then becomes to utilize data on infected and recovered individuals over time to detect both the change points *t_j_*s, as well as the model parameters *β_j_* and *γ_j_* within each fixed segment. The key steps in our proposed change point detection strategy are outlined in Algorithm 1.

Next, we elaborate on the key steps of the Algorithm. First, few notations are defined.

*Notation*. Denote the indicator function of a subset A as l*_A_*. R denotes the set of real numbers. For any vector *v* ∈ ℝ*_p_*, we use ‖*υ*‖_1_, ‖*υ*‖_2_ and ‖*υ*‖_∞_ to denote 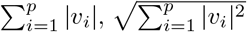 and max_1≤_*_i_*_≤_*_p_* |*υ_i_*|, respectively. The transpose of a matrix ***A*** is denoted by ***A′***.

*Block Fused Lasso:* the objective is to partition the observations into blocks of size *b_n_*, wherein the model parameter *B_t_* remains fixed within each block and select only those blocks for which the corresponding change in the parameter vector is much larger than the others. Specifically, let *n* = *T* − 1 be the number of the times points for the response data *Y_t_* and define a sequence of time points 1 = *r*_0_ <*r*_1_ < ··· <*r_k_*_n_ = *n* + 1 for block segmentation, such that *r_i_* − *r_i_*_−1_ = *b_n_* for *i* =1,…,*k_n_* − 1, 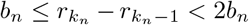, where 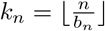 is the total number of blocks. For ease of presentation, it is further assumed that *n* is divisible by *b_n_* such that *r_i_* − *r_i_*_−1_ = *b_n_* for all *i* =1,…,*k_n_*. By partitioning the observations into blocks of size *b_n_* and fixing the model parameters within each block, we set *θ*_1_ = *B*^(1)^ and

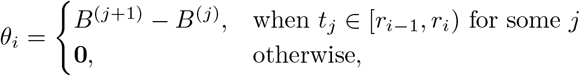

for *i* =2, 3,…,*k_n_*. Note that *θ_i_* ≠ 0 for *i* ≥ 2 implies that *θ_i_* has at least one non-zero entry and hence a change in the parameters. Next, we formulate the following linear regression model in terms of 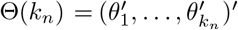:

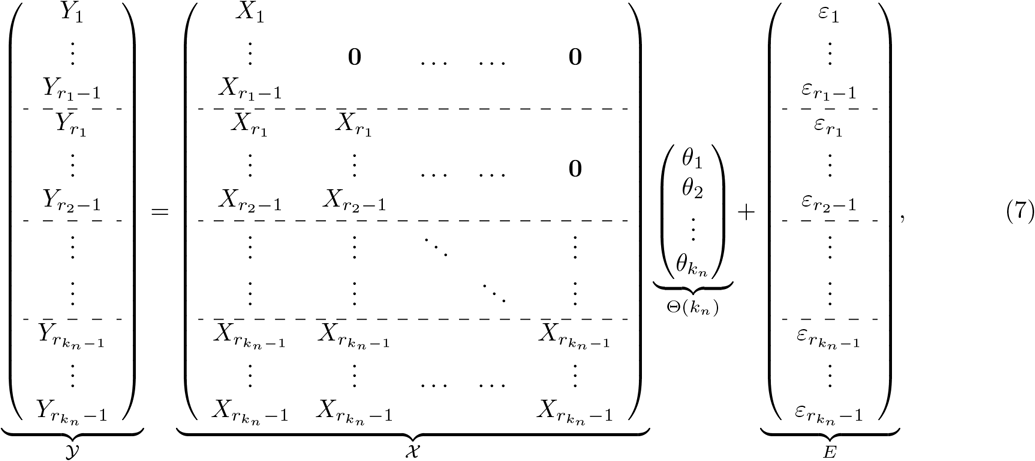

where 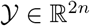, 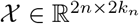, 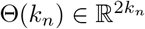 and *E* ∈ ℝ^2^*^n^*.

#### Algorithm 1: Change Point Detection for the Piecewise Stationary SIR model

**Input:** Time series data {*I*(*t*), *R*(*t*)},*t* =1, 2, ···, *T*; regularization parameter *λ_n_*; block size *b_n_* (value of *n* as a function of *T* specified in the sequel).

1. **Block Fused Lasso:** Partition the time points into blocks of size *b_n_* and fix the coefficient parameters within each block. Then, obtain candidate change points 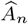 together with estimates of parameters for each block 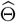 by solving (8).
2. **Hard-thresholding:** For *i* =2, ···, *k_n_*, if 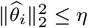, declare that there are no change-points within the *i*-th block; otherwise, select the first time point *r_i_*_−1_ in the *i*-th block as a candidate change point. Denote the set of candidate change points by 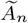.
3. **Block clustering:** Partition the 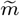 candidate change points 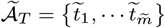 into 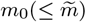 subsets *C* = {*C*_1_,*C*_2_, ···, *Cm*_0_ }, so as to minimize the within-cluster distance.
4. **Exhaustive Search:** Set *l_i_* =(*C_i_* − *b_n_*) and *u_i_* =(*C_i_* + *b_n_*). Apply an exhaustive search method for each time point *s* in the search domain (*l_i_,u_i_*) and estimate the change point 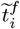 by minimizing the mean square error (MSE) of observations within the interval [*C_i_* − *b_n_,C_i_* + *b_n_*).

**Output:** The estimated parameter matrix 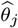 in each segment, 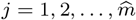 using the block fused lasso. The final estimated change points 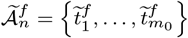.

A simple estimate of parameters Θ(*k_n_*) can be obtained by using an *ℓ*_1_-penalized least squares regression of the form

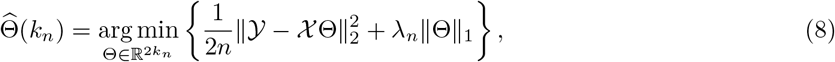

which uses a fused lasso penalty to control the number of change points in the model. This penalty term encourages the parameters across consecutive time blocks to be similar or even identical; hence, only large changes are registered, thus aiding in identifying the change points. Further, a hard-thresholding procedure is added to cluster the jumps into two sets: large and small ones, so that those redundant change points with small changes in the estimated parameters can be removed. We only declare that there is a change point at the end point of a block, when associated with large jump of the model parameters.

*Hard Thresholding:* is based on a data-driven procedure for selecting the threshold *η*. The idea is to combine the *K*-means clustering method (Hartigan and Wong, 1979) with the BIC criterion (Schwarz et al., 1978) to cluster the changes in the parameter matrix into two subgroups. The main steps are:

- Step 1 (initial state): Denote the jumps for each block by 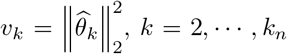 and let *v*_1_ = 0. Denote the set of selected blocks with large jumps as *J* (initially, this is an empty set) and set BIC*^old^* = ∞
- Step 2 (recursion state): Apply *K*-means clustering to the jump vector 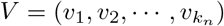 with two centers. Denote the sub-vector with a smaller center as the small subgroup, *V_S_*, and the other sub-vector as the large subgroup, *V_L_*. Add the corresponding blocks in the large subgroup into *J*. Compute the BIC by using the estimated parameters 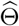 after setting 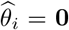 for each block *i* ∉ *J* and denote it by BIC*^new^*. Compute the difference BIC^diff^ = BIC*^new^* − BIC*^old^* and update BIC*^old^* = BIC*^new^*. Repeat this step until BIC^diff^ ≥ 0.

*Block clustering:* the Gap statistic (Tibshirani, Walther and Hastie, 2001) is applied to determine the number of clusters of the candidate change points. The basic idea is to run a clustering method (here, *K*-means is selected) over a grid of possible number of clusters, and to pick the optimal one by comparing the changes in within-cluster dispersion with that expected under an appropriate reference null distribution (for more details, see Section 3 in Tibshirani, Walther and Hastie (2001)).

*Exhaustive search:* Define 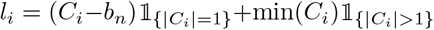 and 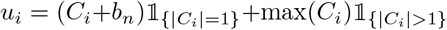, where *C_i_*’s are the subsets of candidates blocks by block clustering procedure. Denote the subset of corresponding block indices by *J_i_*. Define the following local coefficient parameter estimates:

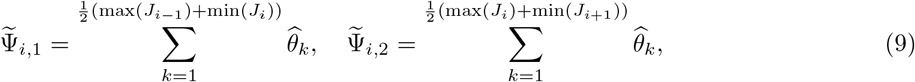

for all *i* =1,…,*m*_0_, where *J*_0_ = {1} and 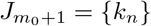.

Now, given a subset *C_i_*, we apply the exhaustive search method for each time point *s* in the interval (*l_i_,u_i_*) to the data set truncated by the two end points in time, min(*C_i_*) − *b_n_* and max(*C_i_*)+ *b_n_*, i.e. only consider the data within the interval [min(*C_i_*)−*b_n_*, max(*C_i_*)+*b_n_*). Specifically, define the final estimated change point 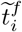 as

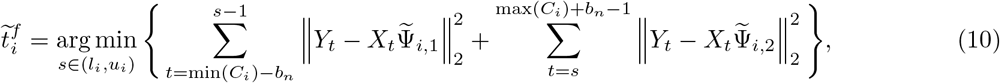

for *i* =1,…,*m*_0_, where 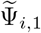 and 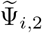 are the local coefficient parameter estimates based on the first step by block fused lasso. Denote the set of final estimated change points from (10) by 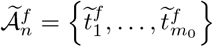.

*Remark:* An alternative approach for detection of break points is to run a full exhaustive search procedure for both single and multiple change point problems. Such procedures are computationally expensive, and not scalable for large data sets. Simple fused lasso is (block of size one) another method, which although computationally fast, it leads to over-estimating the number of break points; hence, it requires additional “screening” steps to remove redundant break points found using the fused lasso algorithm (Harchaoui and Lévy-Leduc, 2010; Safikhani and Shojaie, 2020). Such screening steps usually include tuning several hyper-parameters. This task not only slows down the detection method, but is also not robust. The proposed approach (block fused lasso coupled with hard-thresholding) aims to solve this issue by choosing appropriate block sizes, while it only needs a single tuning parameter (the threshold) to be estimated. *Estimation of Infection and Recovery rates:* Once the locations of break points are obtained, one can estimate the model parameters by running a separate regression for each identified stationary segment of the time series data. The work of Safikhani and Shojaie (2020) shows that this strategy yields consistent model parameter estimates.

*Some Properties of the Detection Procedure:* The estimated change points obtained by the block fused lasso step includes all points with non-zero 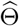, which leads to overestimating the number of the true change points in the model. Hence, the reason for employing a hard-thresholding step to “thin out” redundant change points exhibiting small changes in the estimated coefficients. Nevertheless, the block fused lasso parameter estimates enjoy a prediction consistency property, which implies that the prediction error, i.e. 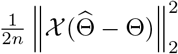, converges to zero with high probability as *n* → +∞. This result is stated and proved in Theorem A.1 in the Appendix A under some mild conditions on the behaviour of the tail distributions of error terms.

After the hard-thresholding step, those candidate change points located far from any true change points will be eliminated when the block size is large enough (*b_n_* = *c* log *n* for a large enough constant *c>* 0). On the other hand, there may be more than one selected change points remaining in the set 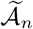 in *b_n_*-neighborhoods of each true change point. The exhaustive search step picks one estimated break point in each such *b_n_*-neighborhoods. Therefore, by selecting *b_n_* = *c* log *n* for a large enough constant *c>* 0, one can conclude that the proposed detection algorithm locates the true break points with an error bounded by the order 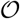. Similar bounds have been found in the literature for similar processes (see for example Chan, Yau and Zhang (2014) for auto-regressive models). For more details, see appendix A.

### 2.2. Model 2: Piecewise Stationary SIR Model with Spatial Heterogeneity

The standard SIR model and its piecewise stationary counterpart do not account for any influence due to inter-region mobility and travel activity. To that end, we extend the previous model *for each region*, by considering the influence exerted by its *q* neighboring regions (cities, counties or states) (see also Srivastava and Prasanna (2020)). Then, for each time point *t* =1, ···, *T* − 1, the model becomes

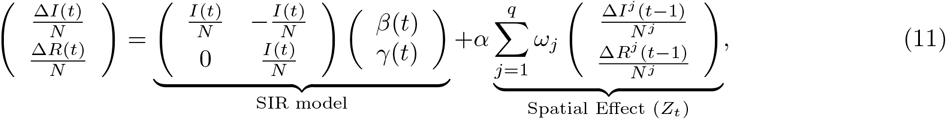

where *N^j^* denotes the total population in neighboring region *j*; *α* is a spatial effect parameter; *ω_j_*’s are spatial weights such that 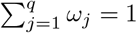. The latter two parameters capture inter-region mobility patterns.

Model (11) can be succinctly written as

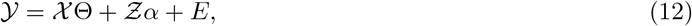

where 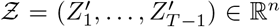 and 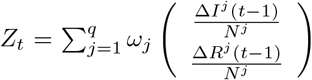 is the weighted average of spatial effect over the neighboring regions at time *t*.

Note that assuming that the influence of the spatial effect component *Z* is small, we can use Model 1 for each region of interest to estimate both the change points and the corresponding transmission and recovery rates. Subsequently, the piecewise constant SIR model, augmented with an overall spatial effect, is used as follows: suppose that there are *m*_0_ change points being detected, denote the final estimated change points by Model 1 as 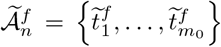. We now formulate the following linear regression model in terms of 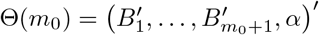:

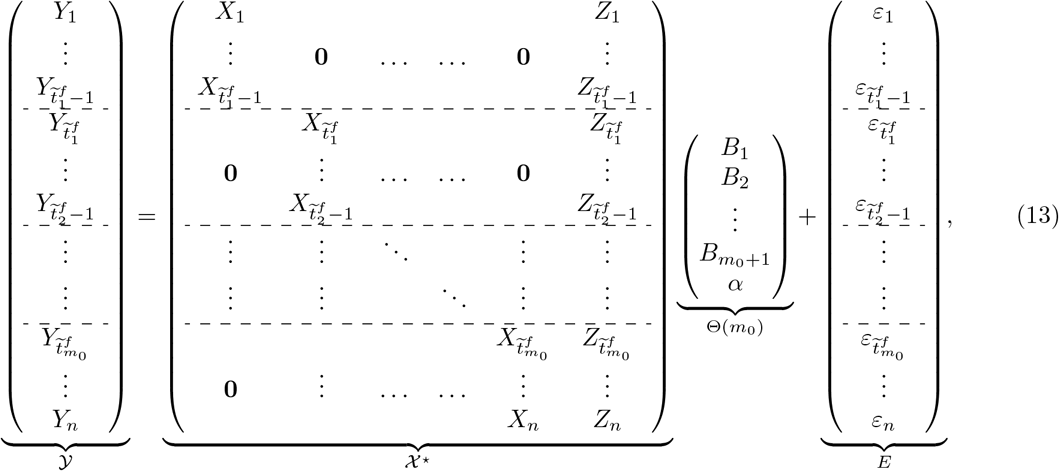

where 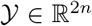, 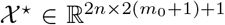, 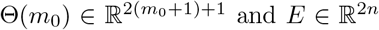 and *E* ∈ ℝ^2^*^n^*. The model parameters are then estimated by least squares.

### 2.3. Model 3: A Stochastic Piecewise Stationary SIR Model with Spatial Heterogeneity

The standard homogeneous SIR model, previously discussed, is deterministic; hence, the output of the model is fully determined by the parameter values of the transmission and recovery rates and the initial conditions. Its stochastic counterpart (Bartlett, 1949; Bailey, 1953), possesses some inherent randomness. The same set of parameter values and initial conditions will lead to an ensemble of different outputs; for an in depth discussion and survey of deterministic and stochastic epidemic models, see Daley and Gani (2001).

The transition probabilities associated with the Markov process underlying the stochastic version of the SIR model are given by

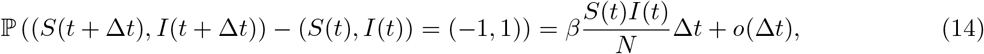

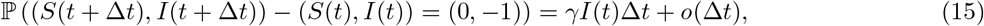

where *S*(*t*),*I*(*t*) ∈{0, 1, 2, ···, *N*} are discrete random variables, denoting the number of susceptible and infected individuals at time *t*.

Using the approximation result in Greenwood and Gordillo (2009) (see also Allen (2017); Buckingham-Jeffery, Isham and House (2018)), the general stochastic epidemic model can be approximated by the stochastic differential equation:

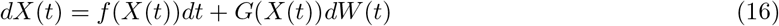

where the random variables *S*(*t*) and *I*(*t*) are continuous,

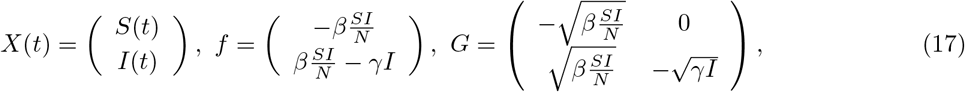

and *W* =(*W*_1_,*W*_2_)′ is a vector of two independent Wiener processes, i.e., 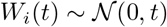.

Under the assumption that *S*(*t*) ≈ *N* and given (16), the stochastic SIR model can be written as:

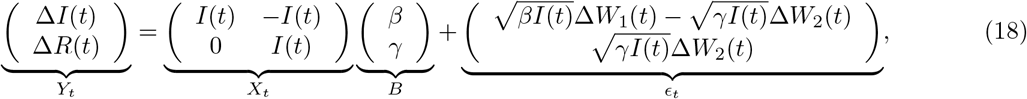

where the increments Δ*W*_1_(*t*) and Δ*W*_2_(*t*) are two independent normal random variables, i.e., 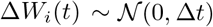.

It can be seen that the resulting regression model, based on the discrete analogue of (18), will have an error term exhibiting temporal correlation, driven by *I*(*t*) and *R*(*t*). An examination of the temporal correlation patterns in the residuals of model 2 from COVID-19 data (see left two panels in Figures 8 and 11 in Appendices B and C) supports this finding. To that end, we model the error process as a Vector Auto-Regressive (VAR) with lag *p*. The corresponding temporal correlation plots for the residuals after inclusion of the VAR(*p*) component are depicted in the right two panels in Figures 8 and 11 in Appendices B and C and clearly show the importance of considering such an error structure.

The piecewise stationary SIR model with spatial heterogeneity and a VAR(*p*) error process is given by

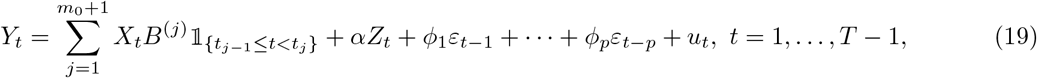

where *u_t_* is white noise with mean 0 and variance *σ*^2^, *ϕ*_1_;…;*ϕ_p_* are the corresponding autoregressive parameters.

The estimation of the model parameters is accomplished in the following three steps: Step 1: Fit model 1 for each region of interest to obtain the change points; Step 2: Obtain the transmission and recovery rates and spatial effect as in Model 2. Step 3: Compute the residuals 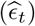 from Step 2 and fit a VAR model to them, see Lütkepohl (2005).

## 3. Simulation Studies

We evaluate the performance of the proposed models on their predictive ability, change point detection and parameter estimation. We consider three simulation scenarios. The details of the simulation settings for each scenario are explained in Section 3.1. All results are averaged over 100 random replicates.

We assess the results for the three models presented: Model 1, the piecewise stationary SIR model; Model 2, the piecewise stationary SIR model with spatial effect; and Model 3, the piecewise stationary SIR model with spatial effect and a VAR(*p*) error process. The out-of-sample mean relative prediction error (MRPE) is used as the performance criterion defined as:

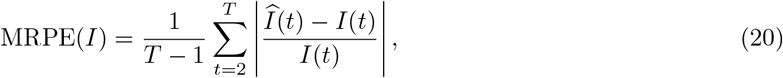

where 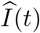 is the predicted count of infected cases at time *t*, and *I*(*t*) the observed one. The MRPE of *R*(*t*) can be obtained by respectively replacing the 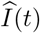 and *I*(*t*) with 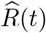 and *R*(*t*). The estimated number of infected cases and recovered cases are defined as

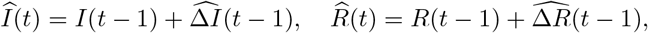

for all *t* =2, ···, *T*. For change point detection, we report the locations of the estimated change points and the percentage of replicates that correctly identifies the change point. This percentage is calculated as the proportion of replicates, where the estimated change points are close to each of the true break points. Specifically, to compute the selection rate, a selected break point is counted as a “success” for the first true break point, *t*_1_, if it falls in the interval 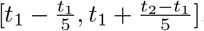. The change point detection results are reported in Table 1. We also evaluate the performance of SIR model with spatial effects by reporting the mean and standard deviation of estimated parameters. The results are reported in Table 2.

**Table 1.** Results of the mean and standard deviation of estimated change point location and the selection rate under the piecewise constant setting.

|  | Model | change point | truth | mean | std | selection rate |
| --- | --- | --- | --- | --- | --- | --- |
| Scenario A | model 1 ($b_n = 4$) | 1 | 0.4 | 0.4012 | 0.0096 | 0.98 |
| | model 1 ($b_n = 4$) | 2 | 0.8 | 0.8 | 0 | 0.97 |
| | model 1 ($b_n = 8$) | 1 | 0.4 | 0.4 | 4e-04 | 0.99 |
| | model 1 ($b_n = 8$) | 2 | 0.8 | 0.8003 | 0.0032 | 0.99 |
| | model 1 ($b_n = 12$) | 1 | 0.4 | 0.4 | 0 | 1 |
| | model 1 ($b_n = 12$) | 2 | 0.8 | 0.7997 | 0.0032 | 1 |
| Scenario B | model 1 ($b_n = 4$) | 1 | 0.5 | 0.4988 | 0.0039 | 1 |
| | model 1 ($b_n = 8$) | 1 | 0.5 | 0.5 | 0 | 1 |
| | model 1 ($b_n = 12$) | 1 | 0.5 | 0.5 | 0 | 1 |
| Scenario C | model 1 ($b_n = 4$) | 1 | 0.5 | 0.4998 | 0.0018 | 1 |
| | model 1 ($b_n = 8$) | 1 | 0.5 | 0.5 | 0 | 1 |
| | model 1 ($b_n = 12$) | 1 | 0.5 | 0.5 | 0 | 1 |

**Table 2.**
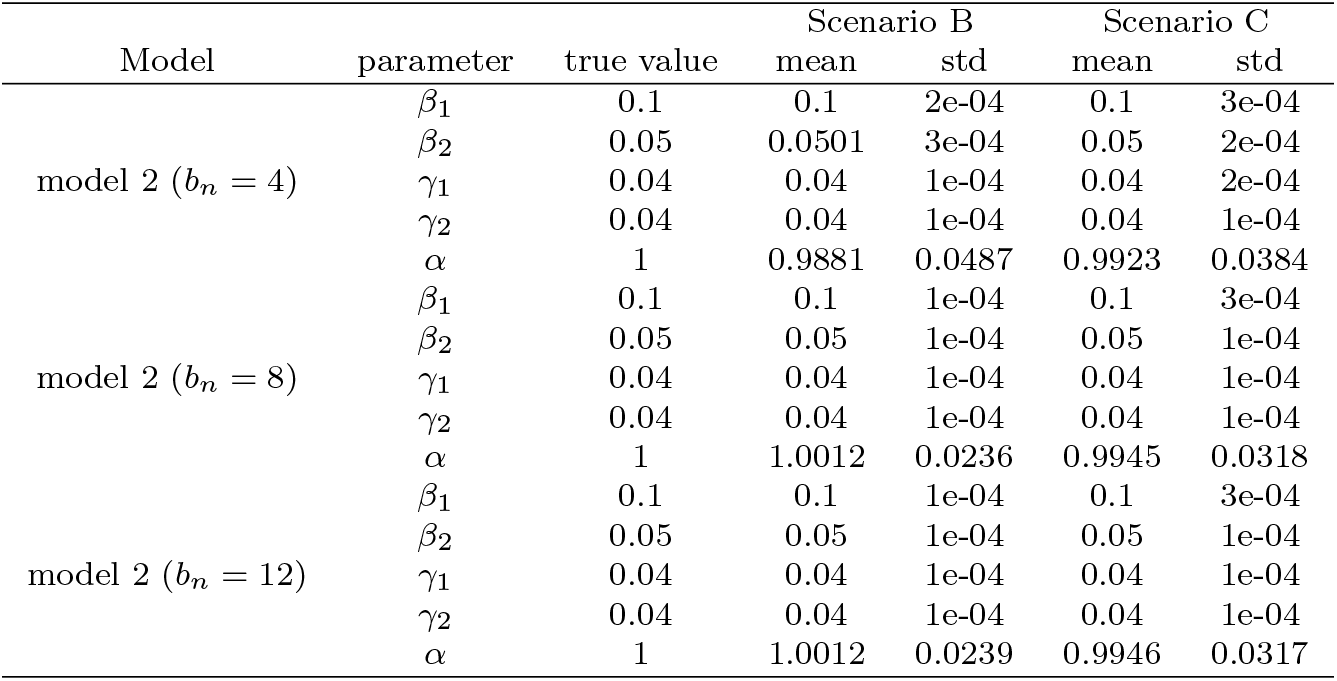
Results of the mean and standard deviation of estimated parameters 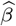, 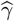, 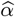.

| Model | parameter | true value | Scenario B |  | Scenario C |  |
| --- | --- | --- | --- | --- | --- | --- |
|  |  |  | mean | std | mean | std |
| model 2 ($b_n = 4$) | $\beta_1$ | 0.1 | 0.1 | 2e-04 | 0.1 | 3e-04 |
| | $\beta_2$ | 0.05 | 0.0501 | 3e-04 | 0.05 | 2e-04 |
| | $\gamma_1$ | 0.04 | 0.04 | 1e-04 | 0.04 | 2e-04 |
| | $\gamma_2$ | 0.04 | 0.04 | 1e-04 | 0.04 | 1e-04 |
| | $\alpha$ | 1 | 0.9881 | 0.0487 | 0.9923 | 0.0384 |
| model 2 ($b_n = 8$) | $\beta_1$ | 0.1 | 0.1 | 1e-04 | 0.1 | 3e-04 |
| | $\beta_2$ | 0.05 | 0.05 | 1e-04 | 0.05 | 1e-04 |
| | $\gamma_1$ | 0.04 | 0.04 | 1e-04 | 0.04 | 1e-04 |
| | $\gamma_2$ | 0.04 | 0.04 | 1e-04 | 0.04 | 1e-04 |
| | $\alpha$ | 1 | 1.0012 | 0.0236 | 0.9945 | 0.0318 |
| model 2 ($b_n = 12$) | $\beta_1$ | 0.1 | 0.1 | 1e-04 | 0.1 | 3e-04 |
| | $\beta_2$ | 0.05 | 0.05 | 1e-04 | 0.05 | 1e-04 |
| | $\gamma_1$ | 0.04 | 0.04 | 1e-04 | 0.04 | 1e-04 |
| | $\gamma_2$ | 0.04 | 0.04 | 1e-04 | 0.04 | 1e-04 |
| | $\alpha$ | 1 | 1.0012 | 0.0239 | 0.9946 | 0.0317 |

### 3.1. Simulation Scenarios

We consider three different simulation settings. In each simulation scenario, we consider three different block size settings. In all scenarios, the block sizes are selected to be *b_n_* =4, 8, 12.

*Simulation Scenario A (SIR model with piecewise constant rates):* the transmission and recovery rates are chosen to be piecewise constant. In this scenario, we set the number of time points *T* = 250, *m*_0_ = 2, the change point *t*_1_ = 100 and *t*_2_ = 200. We choose *β*^(1)^ =0.10, *β*^(2)^ =0.05, *β*^(3)^ =0.01,*γ*^(1)^ =0.04, *γ*^(2)^ = 0.06, *γ*^(3)^ =0.04. Results are based on data generated from the SIR model in (4) with *β*(*t*) ∼ 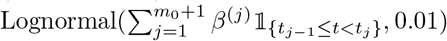 and 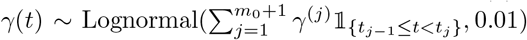. The coefficients for the SIR model are depicted in Figure 1.

**Fig 1:**
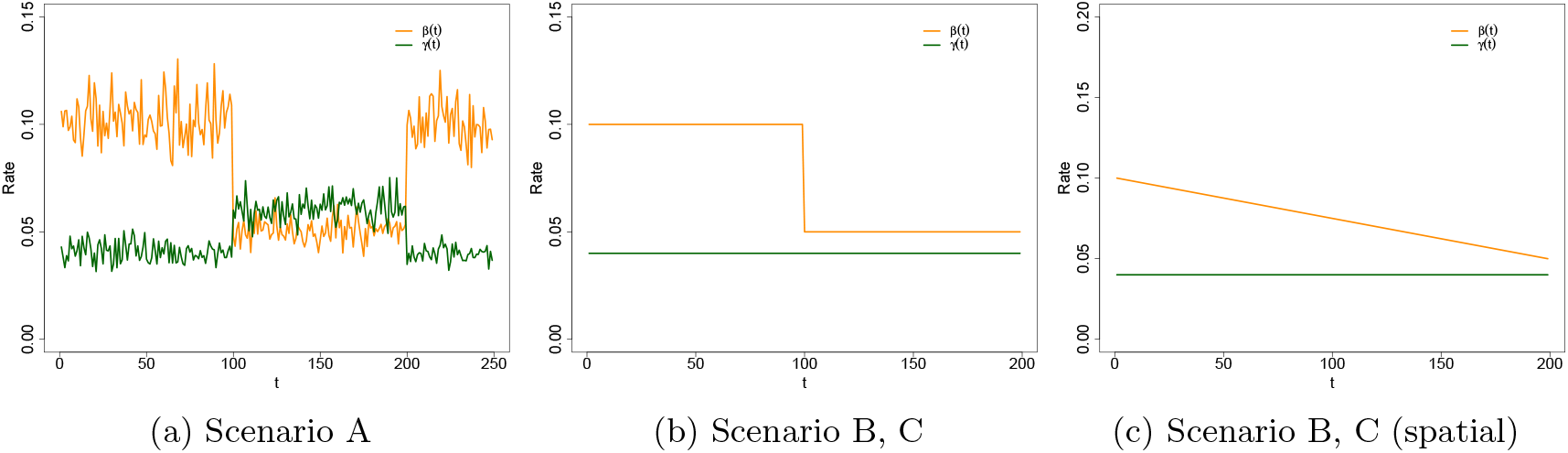
True transmission rate and recovery rate in Scenario A, B, C. The left plot provides the true transmission rate *β*(*t*) and recovery rate *γ*(*t*) in the SIR model in Scenario A; the middle plot provides the true transmission rate *β*(*t*) and recovery rate *γ*(*t*) in the SIR model in Scenario B and C; the right plot provides the true transmission rate *β^s^*(*t*) and recovery rate *γ^s^*(*t*) for generating the spatial component in Scenario B and C.

*Simulation Scenario B (SIR model with piecewise constant rates and spatial effect):* the transmission and recovery rates are chosen to be piecewise constant. In this scenario, we set the number of time points *T* = 200, *m*_0_ = 1, the change point 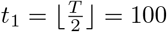. We choose *α* = 1, *β*^(1)^ =0.10, *β*^(2)^ =0.05, *γ*^(1)^ =0.04, *γ*^(2)^ =0.04, 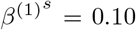, 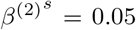, 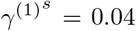, 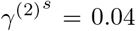. We construct a sequence of *T* − 1 values of 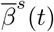 decreasing from 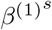 to 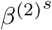and a sequence of *T* − 1 values of 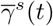 increasing from 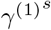 to 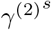 on the linear scale and then generate the spatial effect data from SIR model in (4). By plugging in the spatial effect data, we generate the response variable *Y_t_* from (11) with additional white noise error term from 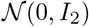. The SIR model’s coefficients for the response variable and the spatial effect variable are depicted in Figure 1. *Simulation Scenario C (piecewise constant SIR model with spatial e*ff*ect and VAR(*1*) structure in error term):* the data are generated based on (19) with piecewise constant transmission and recovery rates. Similar to scenario B, we set the number of time points *T* = 200, *m*_0_ = 1, the change point 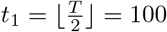, *β*^(1)^ =0.10, *β*^(2)^ = 0.05, *γ*^(1)^ =0.04 and *γ*^(2)^ =0.04. For the spatial effect, we set *α* = 1, 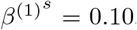, 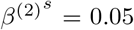, 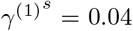, 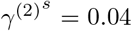. The sequence of *T* − 1 values of 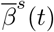 and 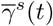 are on the linear scale, same as above in scenario B. We first generate the spatial effect data from SIR model in (4) with parameter 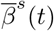 and 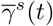 and generate the error term by VAR(1) model with the covariance matrix of the noise process Σ*ε* =0.1*I*_2_. By plugging in the spatial effect data and error term data, we generate the dataset of the response variable *Y_t_* from (19). The SIR model’s coefficients for the response variable and the spatial effect variable are depicted in Figure 1. The autoregressive coefficient matrix has entries 0.8, 0, 0.2, 0.7 from top left to bottom right.

### 3.2. Simulation Results

The mean and standard deviation of the location of the selected change point, relative to the the number of time points *T* – i.e., 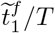 – for all simulation scenarios are summarized in Table 1. The results clearly indicate that, in the piecewise constant setting, our procedure accurately detects the location of change points. The results of the estimated transmission rate 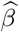, recovery rate 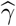 and spatial effect 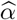 in Table 2 also suggest that our procedure produces accurate estimates of the parameters, under the various settings considered.

In Scenario C, we generate additional 20 days worth of data for prediction testing. The results of the out-of-sample mean relative prediction error (MRPE) for *I*(*t*) and *R*(*t*) are provided in Table 3. The results indicate that adding the spatial effect can significantly improve the prediction, when the spatial component influences the individual data series.

**Table 3.** Results of out-of-sample mean relative prediction error (MRPE) in simulation scenarios C. IR includes both I(t) and R(t)

| Scenario C | Model | MRPE(IR) | MRPE($I(t)$) | MRPE($R(t)$) |
| --- | --- | --- | --- | --- |
| | model 1 ($b_n = 4$) | 0.000255 | 0.000367 | 0.000142 |
| | model 1 ($b_n = 8$) | 0.000252 | 0.000362 | 0.000142 |
| | model 1 ($b_n = 12$) | 0.000252 | 0.000362 | 0.000142 |
| | model 2 ($b_n = 4$) | 2.9e-05 | 4.5e-05 | 1.3e-05 |
| | model 2 ($b_n = 8$) | 2.6e-05 | 3.9e-05 | 1.3e-05 |
| | model 2 ($b_n = 12$) | 2.6e-05 | 3.9e-05 | 1.3e-05 |
| | model 3 ($b_n = 4$) | 2.8e-05 | 4.3e-05 | 1.3e-05 |
| | model 3 ($b_n = 8$) | 2.4e-05 | 3.7e-05 | 1.2e-05 |
| | model 3 ($b_n = 12$) | 2.4e-05 | 3.7e-05 | 1.2e-05 |

The estimated transition matrices of the VAR component are close to the true values, regardless of block size selection. For example, for *b_n_* = 4, the entries of estimated autoregressive coefficients matrix which are averaged among 100 replicates are 0.7577, -0.0057, 0.2361, 0.6663, with standard deviations 0.1125, 0.0461, 0.3582, 0.1147, respectively, while the entries of true autoregressive coefficients matrices are 0.8, 0, 0.2, 0.7 from top left to bottom right. Moreover, for *b_n_* = 8, the entries of estimated autoregressive coefficients matrix are 0.7652 -0.0057, 0.2088, 0.6767, with standard deviations 0.0533, 0.0464, 0.0459, 0.0573, respectively. These results confirm the good performance of the proposed algorithm in finite sample while also indicates its robustness with respect to block size selection.

## 4. Application to State and County Level COVID-19 Data

### 4.1. Data Description

The COVID-19 data used in this study are obtained from The New York Times (2020). The curated data and code used in the analysis are available at the authors’ GitHub repository^1^. The analysis is performed both at the state and county level and the raw data include both cases and deaths, as reported by state and local health department and organized by the NY Times. However, due to lack of complete information on recovered individuals (which are an important covariate in the models considered but the daily number of recovered cases are only reported at the national level (Wang et al., 2020b)), we calculate the number of recovered cases for each region (state/county) as follows: death in the region, multiplied by the nationwide cumulative recovered cases and divided by the nationwide deaths. Specifically, here we assume that the recovery versus deceased ratio for each state/county is fixed, and is approximated by the nationwide recovery to-death ratio. As coronavirus infections keep multiplied and laboratory testing is limited, reporting only confirmed cases and deaths leads to inconsistent data collection and undercounts of the disease’s impact. On April 14, 2020, CDC advised states to count both confirmed and probable cases and deaths. As more states and localities have started to include probable cases and deaths in their counts, in this study, we rely on the combined cases, which include both confirmed and probable cases. The populations of states and counties are obtained from the U.S. Census Bureau (2019). Further, to decide which neighboring states/counties to include in Model 2, their distance to the target state/county of interest is used. The latter is obtained from National Bureau of Economic Research (2010). Here, we define the regions within 500 miles for states and 100 miles for counties/cities as neighboring regions in model 2 and 3. For those areas with a large number of neighboring regions, such as New York state, New York City and King county, we only consider the top five regions with the smallest distances.

We analyzed the daily count of COVID-19 cases at the state-level from March 1, 2020, to August 18, 2020, and at the county-level from the first day after March 1, 2020, when the region records at least one positive COVID-19 case to August 18, 2020. In particular, for the five states presented next (NY, OR, FL, CA, TX), New York city and King county, *n* = 170; for Miami-Dade county, *n* = 160; for Orleans parish, *n* = 161; for Jefferson parish, *n* = 162; for Charleston, Greenville, Richland and Horry counties, *n* = 165, 156, 154, 156, respectively.

Most of the states together with New York City, King and Miami-Dade counties were selected due to being severely affected for a period of time during the course of COVID-19. The remaining counties illustrate interesting patterns gleaned from the proposed models.

Let *I*(*t*) and *R*(*t*) denote the number of infected and recovered individuals (cases) on day *t*. Day 1 refers to the first day after March for which the region records at least one positive COVID-19 case. Figure 2 depicts the actual case numbers *I*(*t*) and *R*(*t*) in the five states considered.

**Fig 2:**
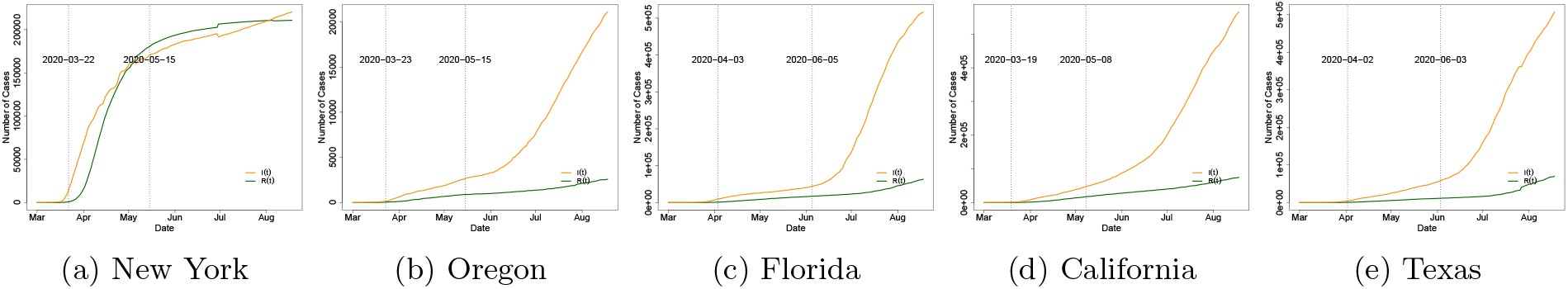
Number of infected cases (dark orange) and recovered cases (dark green) in several states. The first vertical black dotted line indicates the “stay-at-home” order start date for each state, while the second vertical black dotted line indicates the “reopening” start date.

### 4.2. Results for US States

The various models considered are applied on scaled versions (divided by their standard deviations) of the predictors matrix *X_t_* and the response vector *Y_t_*. Further, the block size is set to *b_n_* = 7, i.e. partition the observations into blocks of 7 days for all regions except Texas, wherein a block size *b_n_* = 5 is used. We choose different block sizes since, in some regions, there are more than one break points of the transmission rate very close to each other which violates the assumptions of our model. To mitigate this issue, we select a smaller block size to detect most of the change points. Another situation is that the transmission rate in some regions exhibit many small noise disturbances. In this case, we use a larger block size to smooth out the parameter estimates and detect the change point more accurately. Finally, the tuning parameter λ*_n_* is selected via a cross-validated grid search (Hastie, Tibshirani and Friedman, 2009).

For Model 2, we consider four different types of weights: equal weights (Model 2.1), distance-based weights (Model 2.2), similarity-based weights (Models 2.3 and 2.4). In Model 2.1 and 2.2, the neighboring regions are selected based on distance. For states, a threshold of 500 miles is used and the resulting neighbors are displayed in Table 4. When the spatial resolution is high (county level aggregated data), neighboring regions may exhibit similar patterns in terms of the evolution of transmission and recovery rates. Therefore, constructing weight matrices based on distance is meaningful as it is a common practice in spatial statistics (Cressie, 2015). Such spatial smoothing through proper weight matrices is specially helpful in increasing the statistical power through increasing the sample sizes, hence yielding to more accurate predictions.

**Table 4.** Neighboring states by distance (for model 2.1 and model 2.2).

| Region | Neighboring Regions (states: within 500 miles) |
| --- | --- |
| New York | Connecticut, New Hampshire, New Jersey, Pennsylvania, Vermont |
| Oregon | California, Idaho, Nevada, Washington |
| Florida | Alabama, Georgia, South Carolina |
| California | Arizona, Nevada, Oregon, Utah |
| Texas | Arkansas, Kansas, Louisiana, New Mexico, Oklahoma |

However, the evolution of COVID-19 may exhibit different patterns across neighboring states due to the coarse spatial resolution. Thus, defining weight matrices based on distance may not be ideal. Hence, Models 2.3 and 2.4 select regions based on similarities of infected/recovered cases. Similarity between the region of interest and the *j*-th potential similar region is defined as

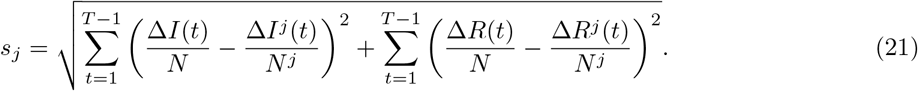

Model 2.3 uses the top five regions with the smallest similarity score, while Model 2.4 uses all states in the country. For states, the resulting neighbors for Model 2.3 are displayed in Table 5.

**Table 5.** Neighboring states by similarity score (for model 2.3).

| Region | Neighboring Regions (states: selected from all states in the country) |
| --- | --- |
| New York | Massachusetts, Connecticut, Michigan, Pennsylvania, District of Columbia |
| Oregon | West Virginia, Wyoming, Montana, Ohio, Washington |
| Florida | South Carolina, Nevada, Alabama, Georgia, Mississippi |
| California | North Carolina, Utah, Oklahoma, Idaho, Missouri |
| Texas | Idaho, Georgia, California, Nevada, South Carolina |

In the equal weight setting (Model 2.1), *ω_j_* =1*/q* for any *j* =1,…,*q*. In both the distance-based weight and similarity-based weight settings, power distance weights are used, wherein the weight is a function of the distance/similarity to the neighboring region:

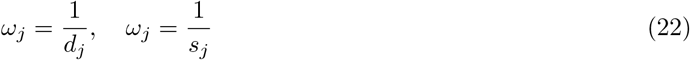

where *d_j_* is the distance score and *s_j_* is the similarity score for the *j*-th region. Under the constraint that 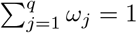, we obtain the normalized weights as:

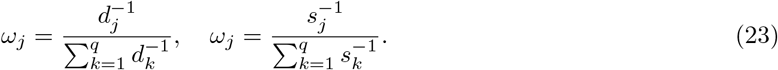

Before applying Model 3, we compare the in-sample and out-of-sample MRPEs (defined in Section 3) in all four variants of Model 2 and select the parameter values estimated by the best-performing model. In subsequent analysis, the results from Model 2.3 are reported, since it proved to be the best performing one.

As expected, change points detected for state data are related to “stay-at-home” orders, or phased reopening dates issued by state governments. We define the reopening date as the time when either the “stay-at-home” order expired or state governments explicitly lifted orders and allowed (select or even all) businesses to reopen (Mervosh et al., 2020). The “stay-at-home” and reopening dates for each state are shown in Table 6. In Model 1, a change point is detected from March to April for four states: New York, Oregon, Florida, and California. These change points coincide with the onset of “stay-at-home” orders and correspond to a significant decrease in the transmission rate. The first change point detected for Florida is around two weeks after the state’s Governor signed a statewide “stay-at-home” order, which is consistent with the fact that COVID-19 symptoms develop 2 days to 2 weeks following exposure to the virus. The first change points in New York and Oregon are detected around 20 days after the statewide “stay-at-home” order, which is longer than Florida. This delayed change point is probably because New York was a hot spot at that time since the virus was in circulation before the start of our observation period (March 1), and thus it took a much longer time to suppress the transmission rate by the statewide “stay-at-home” order. On the other hand, the death toll in New York, which contributes to the recovery rate, continued to rise from the end of March to the beginning of April, even though the “stay-at-home” policy was in effect. Although Texas exhibits a clear downward trend of transmission in April, as gleaned from the estimated transmission rate *β*(*t*) shown in the Figure 7, Model 1 fails to detect the change point for Texas in April. This is mainly because its transmission rate keeps decreasing smoothly during the month of May and violates the model’s piecewise constant assumptions.

**Table 6.** Statewide “Stay-at home” plan and reopening plan begin dates, along with the detected change points (CPs) in five states.

|  | New York | Oregon | Florida | California | Texas |
| --- | --- | --- | --- | --- | --- |
| “Stay-at home” plan | March 22 | March 23 | April 3 | March 19 | April 2 |
| Detected CP | April 11 | April 11 | April 18 | April 29 | May 15 |
| Reopening plan | June 8 (Phase 1)<br>June 22 (Phase 2)<br>July 6 (Phase 3) | May 15 (Phase 1) | May 4 (Phase 1)<br>June 5 (Phase 2) | May 8 (Phase 1)<br>June 12 (Phase 2) | April 30 (Phase 1)<br>May 22 (Phase 2)<br>June 3 (Phase 3) |
| Detected CP | - | May 15 | July 18 | - | June 15, July 17 |

\*NY reopening dates vary in counties. Here, we use the NYC reopening dates instead.

*Remark:* The daily new cases data in California and Texas are highly skewed. This could be another explanation on why the detected change points by our algorithm are not close enough to the “stay-at-home” order start date. To remedy this issue, we considered applying log transformation to the data for these two states to deal with the skewness. The new break points detected by our method applied to the transformed data are much closer to the “stay-at-home” order start date. Specifically, for California, the detected change point occurs at April 8th -almost two weeks after the “stay-at-home” order start date-while the detected change point for Texas occurs at April 10th (almost a week after the “stay-at-home” order start date). A more detailed summary of these findings are stated in Section 5 of the supplementary materials.

As can be seen from Figure 7, in addition to the downward trend after lockdowns have been put in place, Florida and Texas have a clear upward trend after state reopenings began in May. Two change points in June and July are detected for Texas and Florida, which relate to their reopening. Note that in Florida, there is another jump in estimated parameters in June 21st. However, since this jump is close to the already selected change point in July, the algorithm does not pick this time point as a new break point since it violates the modeling assumption that true break points can not be close to each other. Such assumptions are quite natural and common in the anomaly detection literature (Safikhani and Shojaie, 2020).

Note that the restriction in either phase 2 reopening plan in Florida or the phase 3 reopening plan in Texas is quite similar in terms of restaurants, bars, and entertainment businesses. Starting June 5, restaurants and bars in Florida could increase their indoor seating to 50% capacity. Movie theaters, concert venues, arcades, and other entertainment businesses can also open at 50% capacity. Starting June 3, all businesses in Texas could expand their occupancy to 50% with certain exceptions. Bars may increase their capacity to 50% as long as patrons are seated.

Assuming that Florida had not begun the phase 2 reopening plan on June 5, our model predicts 50,585 infected cases by June 12 while the actual number of infected cases is 52,269 (3.3% higher). Similarly, by June 19, our model predicts 57,895 infected cases, while the actual number of infected cases is 69,571 (20.2% higher). Similarly, suppose that Texas had not begun the phase 3 reopening plan on June 3, then by June 10, our model predicts 69,054 infected cases, while the actual number of infected cases is 69,317 (around 0.4% higher). By June 17, our model predicts 82,120 infected cases, while the actual number of infected cases is 85,823 (4.5% higher).

In July, many states paused plans to reopen, amid rising infected case counts. These pausing actions effectively slowed the spread of COVID-19, as can be clearly seen in the downward trend of the transmission rate in July in both Florida and Texas. Hence, a third change point is being detected in Texas and Florida, mainly related to this pausing.

The fitted number of infected cases and recovered cases are defined as

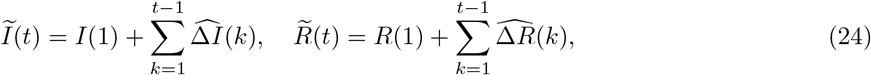

for all *t* =2,…,*T*.

The observed and fitted number of infected cases are displayed in Figure 3. In summary, the piecewise constant SIR model (Model 1) significantly improves the performance in prediction of the number of infected cases compared with the baseline SIR model with fixed rates. Model 2.3 further improves the fit of the data for the California and Texas, due to the addition of a spatial smoothing effect. Finally, Model 3 provides further improvements, in particular for New York and Florida.

To determine the significance level of the spatial effect in model 2, we provide the estimate, p-value, and 95% confidence intervals for the parameter α in Table 15. We find that the influence of the infected or recovered cases in adjacent states is statistically significant (p-value < 0.05) for all states.

**Fig 3:**
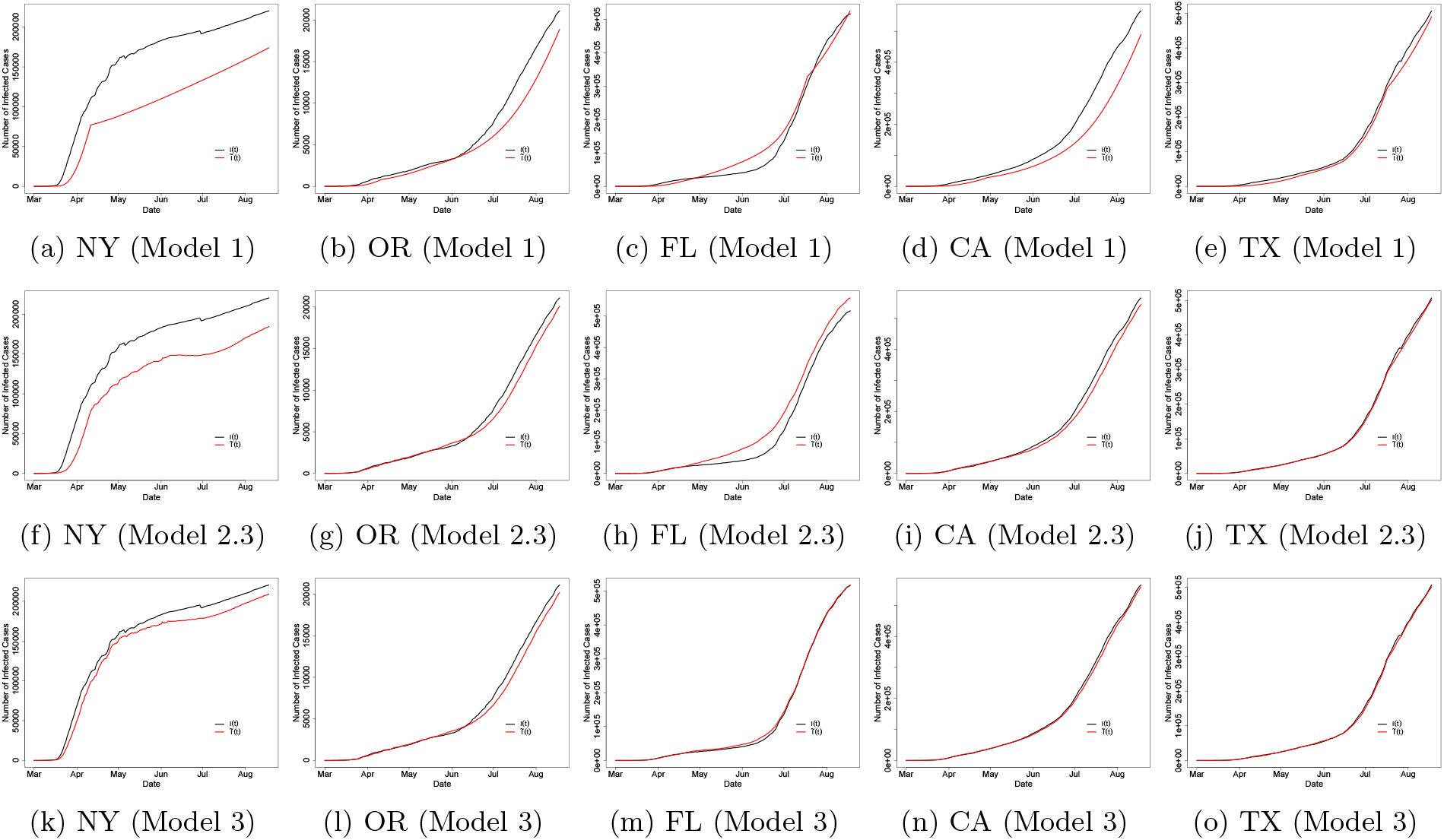
Observed (black) and fitted (red) number of infected cases estimated by three models in five states.

Next, we assess the prediction performance for the three proposed models and the baseline SIR one. The out-of-sample mean relative prediction error (MRPE) is used as the performance, given by

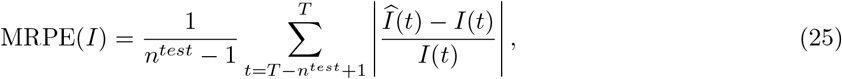

where 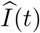 is the predicted number of infected cases at time *t*; and *I*(*t*) is the observed number of infected cases at time *t*. The MRPE of *R*(*t*) can be obtained by respectively replacing the 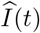 and *I*(*t*) with 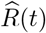 and *R*(*t*). We set the last two weeks of our observation period as the testing period and use the remaining time points for training the model. Note that predicted number of infected and recovered cases are defined as

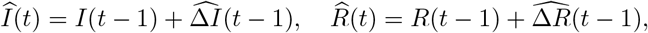

for all *t* =2,…,*T*, where the 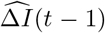 and 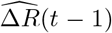 are predicted by using the estimated parameters from the training period of the models.

The results of out-of-sample MRPE of *I*(*t*) and *R*(*t*) in the five states are reported in Tables 7 and 8. The calculated MRPEs of *I*(*t*) show that the model 3 which includes spatial effects and VAR temporal component outperforms the other models. Spatial smoothing itself (model 2) reduces the prediction error significantly in some states. For example, in Florida, the spatial smoothing reduced the MRPE by 80% when using model 2.2 (distanced-based weight) while in Oregon and Texas, the spatial smoothing reduced the MRPE (I) by more than 60% when using model 2.4 and model 2.3, respectively (similarity-based weight). In the state of New York, the prediction error reduction using spatial effects was not significant mainly due to the fact that its neighbors had different COVID-19 progression compared to New York. New York state was the hotspot at the beginning of the pandemic while the neighboring states had a surge in the number of cases much later. Finally, the reduction in MRPEs using model 3 (as reported in Tables 7 and 8) justifies the presence of the VAR component in the modeling framework. Additional results related to the VAR component (including the estimated auto-regressive parameters) are reported in Section 4 of the supplementary materials.

**Table 7.** Out-of-sample mean relative prediction error (MRPE) of I(t).

|  | NY | OR | FL | CA | TX |
| --- | --- | --- | --- | --- | --- |
|  | MRPE(I) | MRPE(I) | MRPE(I) | MRPE(I) | MRPE(I) |
| Model 1 | 0.0017 | 0.0091 | 0.0168 | 0.0076 | 0.0117 |
| Model 2.1 | 0.0015 | 0.0034 | 0.0029 | 0.0029 | 0.0042 |
| Model 2.2 | 0.0015 | 0.0037 | 0.0028 | 0.0032 | 0.0039 |
| Model 2.3 | 0.0014 | 0.0047 | 0.0032 | 0.0029 | 0.0031 |
| Model 2.4 | 0.0021 | 0.0033 | 0.0053 | 0.0032 | 0.004 |
| model 3 | 0.0015 | 0.0046 | 0.0032 | 0.0029 | 0.0032 |

**Table 8.** Out-of-sample mean relative prediction error (MRPE) of R(t).

|  | NY<br>MRPE(R) | OR<br>MRPE(R) | FL<br>MRPE(R) | CA<br>MRPE(R) | TX<br>MRPE(R) |
| --- | --- | --- | --- | --- | --- |
| Model 1 | 0.0054 | 0.0094 | 0.0062 | 0.0054 | 0.0083 |
| Model 2.1 | 0.0019 | 0.0099 | 0.0111 | 0.0052 | 0.0076 |
| Model 2.2 | 0.0018 | 0.0098 | 0.0111 | 0.0061 | 0.0076 |
| Model 2.3 | 7e-04 | 0.009 | 0.0131 | 0.0041 | 0.0067 |
| Model 2.4 | 0.0052 | 0.0086 | 0.0074 | 0.0044 | 0.0068 |
| model 3 | 7e-04 | 0.0089 | 0.0131 | 0.0039 | 0.0067 |

In Figure 4, we provide the predicted values in the last 2 weeks from all three different models in three states: New York, Florida and California. In both the daily infected cases Δ*I*(*t*) and the daily recovered cases Δ*R*(*t*), the predictions by model 2 and model 3 perform better than model 1 in all three states.

**Fig 4:**
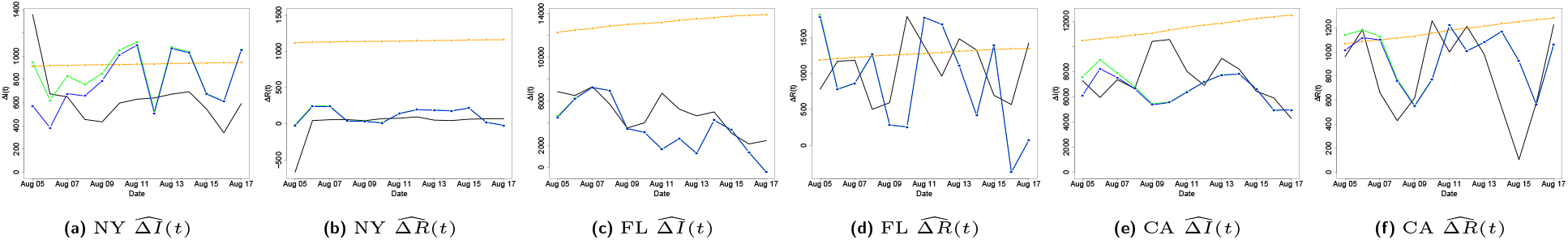
Prediction of response variable *y_h_* in three models. Black line: true value; orange: model 1; green: model 2; blue: model 3.

### 4.3. Results for Selected US Counties

We focus on the following counties in this analysis: New York City, King County in Washington, Miami-Dade County in Florida, Orleans and Jefferson parishes in Louisiana and Charleston, Greenville, Richland, and Horry in South Caroline. Note that The New York Times (2020) reports a single value for New York City which includes the cases in New York, Kings, Queens, Bronx and Richmond Counties.

The block size *b_n_* is set to 7 for New York City, King and the South Carolina counties. For Miami-Dade, a smaller block size *b_n_* = 5 is used, while for Louisiana counties a larger one *b_n_* = 10. Further, for determining their neighbors displayed in Table 9, a threshold of 100 miles is used. Model 2.3 uses the top five counties in the corresponding state with the smallest similarity score, while Model 2.4 uses all counties in the given state. The resulting neighbors for Model 2.3 are displayed in Table 10.

Since the parameters obtained from (5) vary greatly, the 7-day moving average of transmission *β*(*t*) and recovery *γ*(*t*) rates is provided in Figure 9 and 10. As can be seen from those figures, our estimated piecewise parameters are very close to the real parameters in the 7-day moving average.

**Table 9.** Neighboring cities/counties/parishes by distance (for model 2.1 and model 2.2).

| Region | Neighboring Regions (counties: within 100 miles) |
| --- | --- |
| New York City (NY) | Hudson, Essex, Union, Bergen, Nassau |
| King County (WA) | Pierce, Island, Kitsap, Kittitas, Snohomish |
| Miami-Dade County (FL) | Broward, Collier, Hendry, Palm Beach, Monroe |
| Orleans Parish (LA) | Jefferson, St. Charles, St. John the Baptist, St. Tammany, Hancock |
| Jefferson Parish (LA) | Lafourche, Orleans, Plaquemines, St. Charles, Terrebonne |
| Charleston (SC) | Dorchester, Beaufort, Berkeley, Colleton, Georgetown |
| Greenville (SC) | Polk, Henderson, Anderson, Pickens, Spartanburg |
| Richland (SC) | Calhoun, Lexington, Sumter, Fairfield, Kershaw |
| Horry (SC) | Columbus, Dillon, Marion, Florence, Georgetown |

**Table 10.** Neighboring cities/counties/parishes by similarity score (for model 2.3).

| Region | Neighboring Regions (counties: in the same state) |
| --- | --- |
| New York City (NY) | Erie, Dutchess, Monroe, Columbia, Niagara |
| King County (WA) | Thurston, Kitsap, Pierce, Lewis, Grays Harbor |
| Miami-Dade County (FL) | Broward, Osceola, Escambia, Bay, Duval |
| Orleans Parish (LA) | Jefferson, St. Bernard, St. Tammany, St. Charles, Ascension |
| Jefferson Parish (LA) | Orleans, St. Bernard, St. Tammany, Lafourche, St. Charles |
| Charleston (SC) | Horry, Berkeley, Dorchester, Greenville, Orangeburg |
| Greenville (SC) | Pickens, Lexington, Richland, Spartanburg, York |
| Richland (SC) | Lexington, Greenville, Berkeley, Beaufort, York |
| Horry (SC) | Greenville, Berkeley, Charleston, Richland, Pickens |

The statewide and countywide policy start dates and the detected change points are shown in Tables 11 and 12. A change point is detected from March to April for the first three counties. These change points coincide with the onset of “stay-at-home” orders and indicate a significant decrease in the transmission rate. A change point in June is detected in King county and Miami-Dade county, which may be related to the reopening plan after June as both change points correspond to a substantial increase in the transmission rate. Similar to Texas, a third change point is being detected in Miami-Dade county in July due to pausing in the reopening plans.

**Table 11.** Statewide “Stay-at-home” plan and reopening plan begin dates, along with the detected change points in counties/cities/parishes.

|  | New York City | King County | Miami-Dade County | Orleans Parish | Jefferson Parish |
| --- | --- | --- | --- | --- | --- |
| “Stay-at-home” plan (statewide) | March 22 | March 23 | April 3 | March 23 | March 23 |
| Detected change point (model 1) | April 5 | April 12 | April 13 | April 9 | April 9 |
| Reopening plan (statewide) | May 15 | May 30 | June 5 (Phase 2) | June 5 (Phase 2) | June 5 (Phase 2) |
| Detected change point (model 1) | - | June 28 | June 23, July 25 | - | June 27 |

**Table 12.** Statewide “Stay-at-home” plan and reopening plan begin dates, along with countywide mask mandate plan and the detected change points in four counties in South Carolina.

|  | Charleston | Greenville | Richland | Horry |
| --- | --- | --- | --- | --- |
| “Stay-at-home” plan (statewide) | April 7 | April 7 | April 7 | April 7 |
| Reopening plan (statewide) | April 20 | April 20 | April 20 | April 20 |
| Mask mandate (countywide) | July 1 | No mandate | July 6 | July 3 |
| Detected change point (model 1) | June 9, July 6 | May 6, June 20 | May 11, July 24 | July 11 |

Next, using the proposed set of models we discuss two cases that provide interesting insights into policy questions.

*Case I: Neighboring Parishes in Louisiana*. A change point is detected in April for the Orleans and Jefferson parishes in Louisiana. This is primarily due to the fact that the virus circulated widely due to Mardi Gras activities in the end of February, that led to a high transmission rate in the March/early April period. However, the estimated transmission rate is much higher in Orleans, compared to Jefferson, which translates to a much higher number of infections in the former county. After reopening in early June, the transmission rate exhibited an increase (and hence a change point), only for Jefferson county. A possible explanation is that the high transmission rate in March in Orleans county led to a much larger number of infections, thus possibly conferring immunity to a larger portion of the population, while that was not the case in Jefferson county. Hence, in Jefferson’s case, the reopening led to a subsequent increase in the transmission rate. Note that seroprevalence survey data from the CDC^2^ show a much higher percentage of immunity for Louisiana at the time of this writing, but probably not uniformly distributed across the state.

*Case II: Neighboring Counties in South Carolina*. We consider the following four neighboring counties (harleston, Greenville, Richland, and Horry), where mask wearing policies exhibited some variation in their timing, while Greenville county did not impose one. We can see that the transmission rates in these four counties share a very similar pattern with an upward trend in May and a downward trend from late June to August. Although all four counties show a significant decline in transmission rates in July, the mask mandate seems to lead to a reduction in transmission rates. Specifically, in Charleston at the change point in July the estimated transmission rate has a jump of 0.04, which is larger than the one in Greenville’s June change point of 0.02. The difference in jump size may relate to mandating the facial covering. What is difficult to delineate is whether the timing of the imposition of the mask mandate in three out of the four neighboring counties should have occurred earlier, so as the transmission rate not to exhibit such a large jump, especially in the most populous Charleston county.

Figure 11 shows the ACF plot of model 2’s residual time series for three counties. As can be seen from these plots, in Orleans parish, there is a significant temporal dependence among the time-lagged residual time series processes. This temporal dependence has been successfully captured by including the VAR(*p*) model for the residuals, as seen in the ACF plot of the new residuals, i.e. 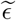.

A more intuitive result is shown in Figure 5, where the fitted values of infected cases by model 2 and model 3 are much closer to the observed value than the predicted value by model 1. Specifically, Model 2.3 improves the fit of the data in all regions, due to the addition of a spatial smoothing effect. Model 3 further improves the fit of the data in New York City and Orleans parish.

**Fig 5:**
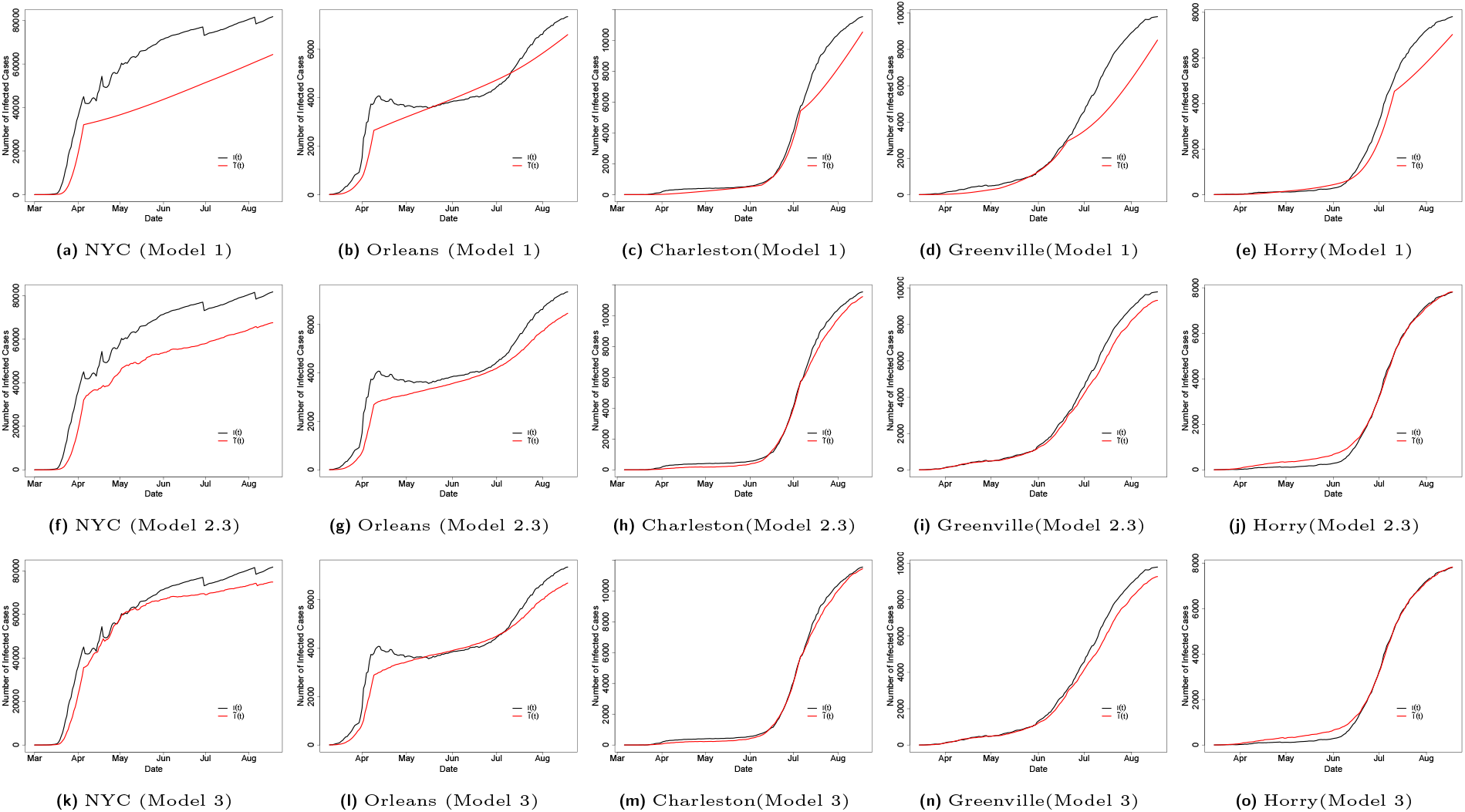
Observed (black) and fitted (red) number of infected cases estimated by three models in parishes/counties/cities.

We also provide the out-of-sample MRPE of *I*(*t*) and *R*(*t*) in Tables 13 and 14. The MRPE of *I*(*t*) results show that the piecewise constant model (model 1) performs the best in New York City, Orleans parish, while the piecewise constant model with spatial effect (model 2) performs the best in King County, Miami-Dade County, Jefferson parish and Richland. Adding the VAR(*p*) (model 3) performs the best in Charleston and Horry. The MRPE of *R*(*t*) results show that the piecewise constant model with spatial effect (model 2) performs the best in King County, Miami-Dade County, Greenville, Richland and Horry, while adding the VAR(*p*) (model 3) performs the best in New York City, Orleans, Jefferson and Charleston.

**Table 13.** Out-of-sample mean relative prediction error (MRPE) of I(t) and R(t).

|  | NYC |  | King |  | Miami-Dade |  | Orleans |  | Jefferson |  |
| --- | --- | --- | --- | --- | --- | --- | --- | --- | --- | --- |
|  | MRPE(I) | MRPE(R) | MRPE(I) | MRPE(R) | MRPE(I) | MRPE(R) | MRPE(I) | MRPE(R) | MRPE(I) | MRPE(R) |
| Model 1 | 0.004 | 0.0063 | 0.0036 | 0.0029 | 0.0169 | 0.008 | 0.003 | 0.0061 | 0.0072 | 0.0069 |
| Model 2.1 | 0.0048 | 0.005 | 0.0037 | 0.0033 | 0.0061 | 0.0068 | 0.0032 | 0.0054 | 0.0065 | 0.0065 |
| Model 2.2 | 0.0043 | 0.005 | 0.0036 | 0.0033 | 0.0053 | 0.0067 | 0.0033 | 0.0054 | 0.0065 | 0.0065 |
| Model 2.3 | 0.005 | 0.0056 | 0.0037 | 0.0032 | 0.0058 | 0.0118 | 0.0033 | 0.0055 | 0.0064 | 0.0063 |
| Model 2.4 | 0.0062 | 0.0064 | 0.0036 | 0.0031 | 0.0157 | 0.0094 | 0.0038 | 0.0062 | 0.0063 | 0.0071 |
| Model 3 | 0.0051 | 0.0047 | 0.0037 | 0.0033 | 0.0058 | 0.0113 | 0.0033 | 0.005 | 0.0065 | 0.0062 |

**Table 14.** Out-of-sample mean relative prediction error (MRPE) of I(t) and R(t).

|  | Charleston |  | Greenville |  | Richland |  | Horry |  |
| --- | --- | --- | --- | --- | --- | --- | --- | --- |
|  | MRPE(I) | MRPE(R) | MRPE(I) | MRPE(R) | MRPE(I) | MRPE(R) | MRPE(I) | MRPE(R) |
| Model 1 | 0.0117 | 0.0195 | 0.0134 | 0.0093 | 0.0143 | 0.0107 | 0.012 | 0.0106 |
| Model 2.1 | 0.0069 | 0.0185 | 0.0085 | 0.0137 | 0.0091 | 0.0125 | 0.0109 | 0.0102 |
| Model 2.2 | 0.0059 | 0.0182 | 0.0078 | 0.0141 | 0.0091 | 0.0127 | 0.0107 | 0.0102 |
| Model 2.3 | 0.0036 | 0.0179 | 0.0041 | 0.0104 | 0.0043 | 0.0094 | 0.004 | 0.0111 |
| Model 2.4 | 0.0063 | 0.0194 | 0.0058 | 0.0127 | 0.0064 | 0.0122 | 0.0083 | 0.0117 |
| Model 3 | 0.0022 | 0.0175 | 0.0041 | 0.0105 | 0.0044 | 0.0095 | 0.0037 | 0.0142 |

Finally, Figure 6 shows the predicted values in the last 2 weeks from all three different models. Here, we provide the prediction plots in three counties: Orleans, Charleston and Richland. In terms of the infected cases, the predictions by model 2 and model 3 perform much better than model 1 in all three counties.

**Fig 6:**
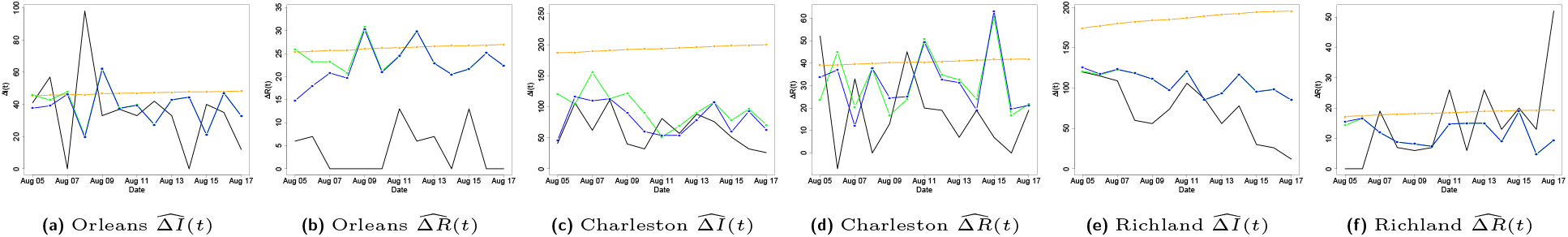
Prediction for the response variable *y_h_* in three models. Black line: true value; orange: model 1; green: model 2; blue: model 3.

## 5. Conclusion and Discussion

COVID-19 has posed a number of challenges for modellers, both due to the lack of adequate data (especially early on in the course of the pandemic) and its characteristics (relative long period before emergence of symptoms compared to SARS and other viruses). A plethora of models -a number of them briefly summarized in the introductory section-were developed, most aiming to provide short and long term predictions of the spread of COVID-19. This work contributes to that goal by developing a piecewise stationary SIR model with neighboring effect and temporal dependence to model the spread of COVID-19 at both state-level and county-level in the United States. Based on the results, a substantial decrease in the transmission rate was detected in the late March/early April period, which is a consequence of many factors, including “stay-at-home” orders, but also change in behavior by individuals once the infectious nature of the disease and the consequences for those severely affected became apparent. Florida and Texas experienced strong jumps in the transmission rates after reopening, which subsequent led to imposition of additional mitigation strategies (mask mandates) and a slower implementation of the multi-phase reopening plans originally devised.

The reasonable short-term forecasts of model 3 (including spatial effects and the VAR component) confirm the existence of spatial and temporal dependence among new daily cases which can not be accounted by the homogeneous deterministic SIR model. Further, the detection of change points in neighboring counties can provide insights into how the spread of COVID-19 impacted different communities at different points in time and also that of mitigation policies adopted by county (state) health adminsitrators.

Our paper did not take the under-reporting or the reported with delay issues of new cases/deaths into account. Therefore, the estimated transmission rates in the model may be over-estimated or under-estimated. Nevertheless, our model helps to understand various trends at the state/countylevels of COVID-19 and also allow researchers to look back at the impact of various mitigation strategies in impacting the course of the epidemic.

## Data Availability

All data used are publicly available and also can be found in the authors' GitHub page with the link below:
https://github.com/ybai69/COVID-19-Change-Point-Detection

## APPENDIX

Some theoretical properties are explained with their proofs in Appendix A. In Appendix B, we provide additional results for U.S. states while in Appendix C, additional results for U.S. counties are stated.

## Appendix A: Theoretical Properties

In this section, we establish the prediction consistency of the estimator from (8). To establish prediction consistency of the procedure, the following assumptions are needed:

(A1.) (Deviation bound) There exist constants *c_i_ >* 0 such that with probability at least 1−*c*_1_exp(−*c*_2_(log 2*n*)), we have

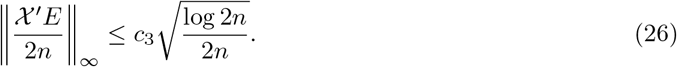
(A2.) There exists a positive constant *M_B_ >* 0 such that

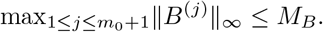

### Theorem A.1

*Suppose A1-A2 hold. Choose* 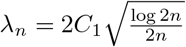 *for some large constant C*_1_ *>* 0, *and assume m*_0_ ≤ *m_n_ with* 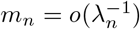*. Then with high probability approaching to 1 and n* → +∞, *the following holds:*

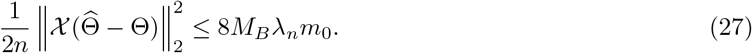

#### Proof of Theorem 1

By the definition of 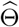 in (8), the value of the function in (8) is minimized at 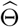. Therefore, we have

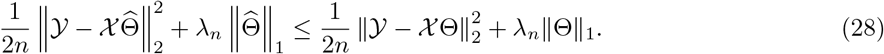

Denoting 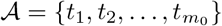 as the set of true change points, we have

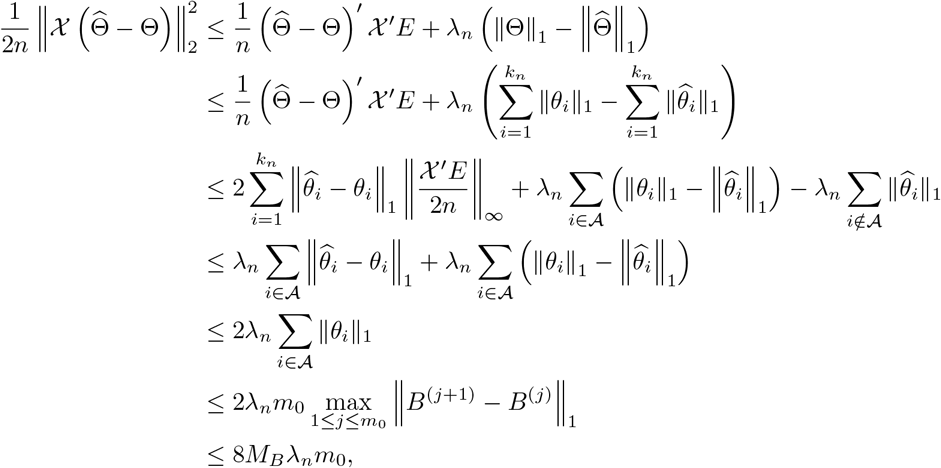

with high probability approaching to deviation bound in (26). This completes the proof. □

Theoretical properties of lasso have been have been studied by several authors (Bickel et al., 2009; van de Geer, Bühlmann and Zhou, 2011; Loh and Wainwright, 2012; Negahban et al., 2012). In controlling the statistical error, a suitable deviation conditions on 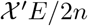 is needed. The deviation bound conditions (e.g. the assumption A1) are known to hold with high probability under several mild conditions. Under the condition that the error term 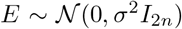, the deviation bound condition holds with high probability by Lemme 3.1 in van de Geer, Bühlmann and Zhou (2011). Given that the *p* (the number of time series components) is small and fixed, we have *n* ≫ log *p*, therefore, in the case where the 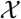 is a zero-mean sub-Gaussian matrix with parameters 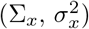, and the error term *E* is a zero-mean sub-Gaussian matrix with parameters 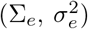, the deviation bound condition holds with high probability by Lemme 14 in Loh and Wainwright (2012).

*Detection Accuracy*. When the block size is large enough, such that log *n/b_n_* remains small, if the selected change point 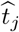 is close to a true change point, the estimated 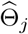 will be large (asymptotically similar to the true jump size in the model parameters); if the selected change point 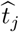 is far away from all the true change points, the estimated 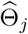 will be quite small (converges to zero as sample size tend to infinity). Therefore, after the hard-thresholding, the candidate change points that are located far from any true change points will be eliminated. In other words, for any selected change point 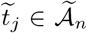, there would exist a true change point 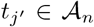 close by, with the distance being at most *b_n_*. Thus, the number of clusters (by radius *b_n_*) seems to be a reasonable estimate for the true number of break points in the model.

On the other hand, since the set of true change points 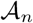 has cardinality less than or equal to the cardinality of the set of selected change points 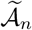, i.e., 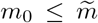, there may be more than one selected change points remaining in the set 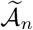 in *b_n_*-neighborhoods of each true change point. For a set *A*, define cluster (*A, x*) to be the minimal partition of *A*, where the diameter for each subset is at most *x*. Denote the subset in cluster 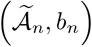 by cluster 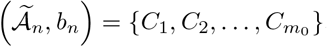, where each subset *C_i_* has a diameter at most *b_n_*, i.e., 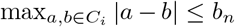. Then with high probability converging to one, the number of subsets in cluster 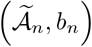 is exactly *m*_0_. All candidate change points in 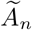 are within a *b_n_*-neighborhood of at least one true change point and therefore, with high probability converging to one, there is a true change point *t_i_* within the interval (*C_i_* − *b_n_,C_i_* + *b_n_*). The distance between the estimated change point and the true change point will be less then 2*b_n_*. Therefore, by selecting *b_n_* = *c* log *n* for a large enough constant *c>* 0, one can conclude that the proposed detection algorithm locates the true break points with an error bounded by the order 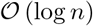.

## Appendix B: Additional Results for U.S. States

**Fig 7:**
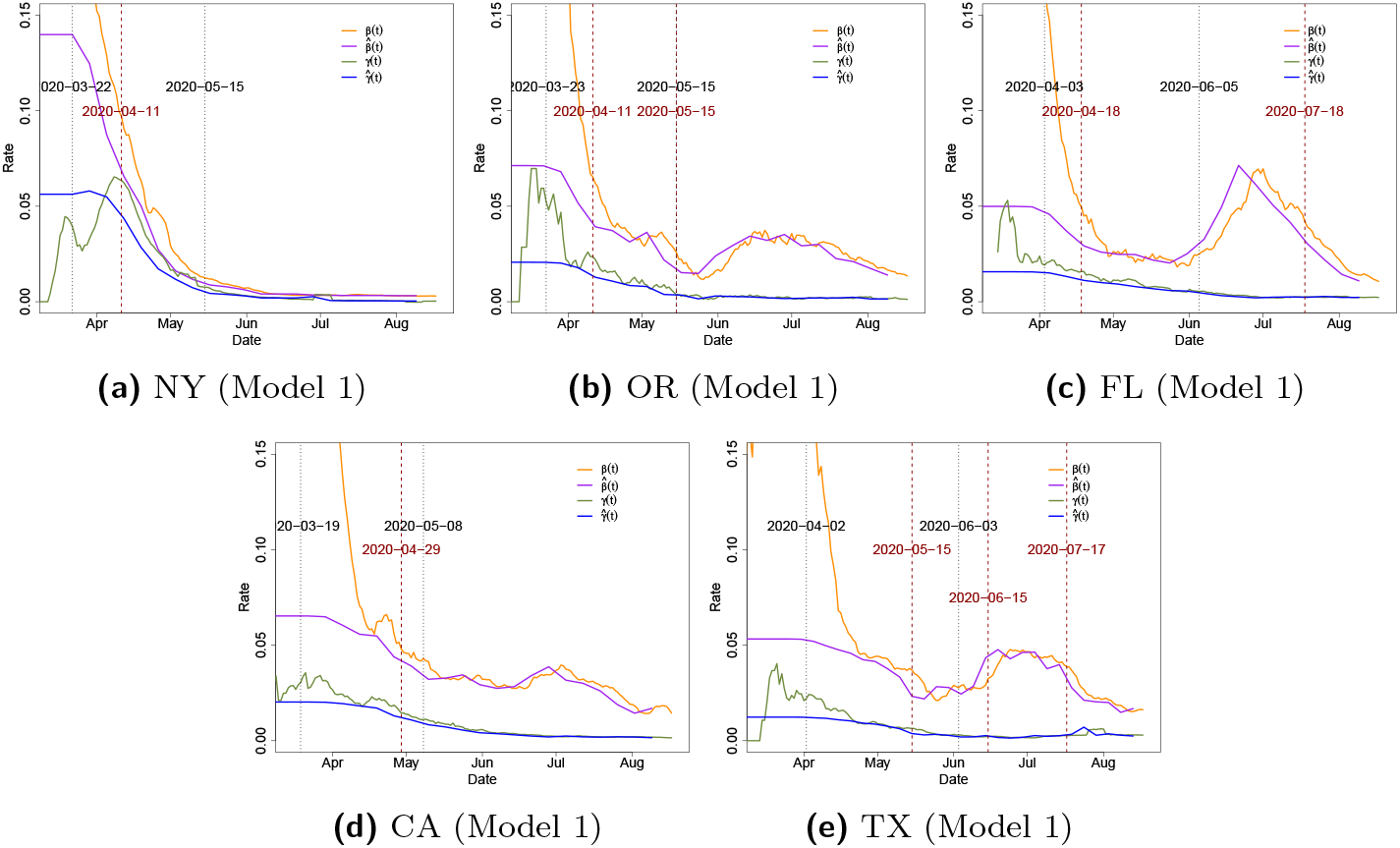
The 7-day moving average of observed transmission rate *β*(*t*) (orange) and recovery rate *γ*(*t*)(green) along with the estimated transmission rate 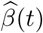 (purple) and recovery rate 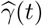 (blue) in several states. The vertical black dotted line indicates the statewide “stay-at-home” order begin date or reopening begin date. The vertical dark red dashed line indicates the estimated change time point for each state.

**Table 15.**
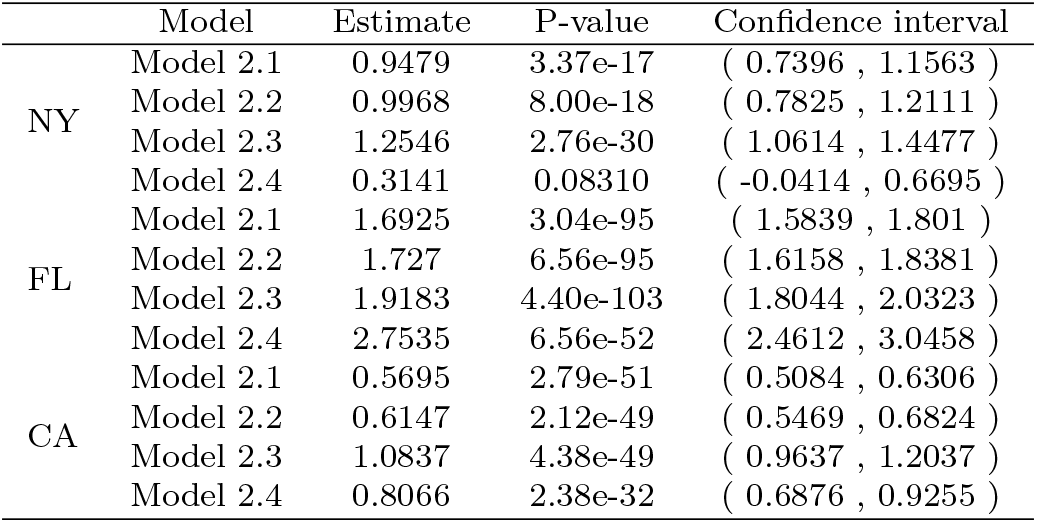
Spatial effect estimation in model 2.

|  | Model | Estimate | P-value | Confidence interval |
| --- | --- | --- | --- | --- |
| NY | Model 2.1 | 0.9479 | 3.37e-17 | ( 0.7396 , 1.1563 ) |
|  | Model 2.2 | 0.9968 | 8.00e-18 | ( 0.7825 , 1.2111 ) |
|  | Model 2.3 | 1.2546 | 2.76e-30 | ( 1.0614 , 1.4477 ) |
|  | Model 2.4 | 0.3141 | 0.08310 | ( -0.0414 , 0.6695 ) |
| FL | Model 2.1 | 1.6925 | 3.04e-95 | ( 1.5839 , 1.801 ) |
|  | Model 2.2 | 1.727 | 6.56e-95 | ( 1.6158 , 1.8381 ) |
|  | Model 2.3 | 1.9183 | 4.40e-103 | ( 1.8044 , 2.0323 ) |
|  | Model 2.4 | 2.7535 | 6.56e-52 | ( 2.4612 , 3.0458 ) |
| CA | Model 2.1 | 0.5695 | 2.79e-51 | ( 0.5084 , 0.6306 ) |
|  | Model 2.2 | 0.6147 | 2.12e-49 | ( 0.5469 , 0.6824 ) |
|  | Model 2.3 | 1.0837 | 4.38e-49 | ( 0.9637 , 1.2037 ) |
|  | Model 2.4 | 0.8066 | 2.38e-32 | ( 0.6876 , 0.9255 ) |

Few additional plots and tables related to the results of applying our method to some U.S. states are provided in this section. In Figure 7, we show the 7-day moving average of measured transmission rate *β*(*t*) and recovery rate *γ*(*t*) by (5), along with the estimated 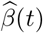 and 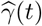 from model 2. We can see that the measured transmission rate *β*(*t*) decreases dramatically during the late March-early April in all five states.

**Fig 8:**
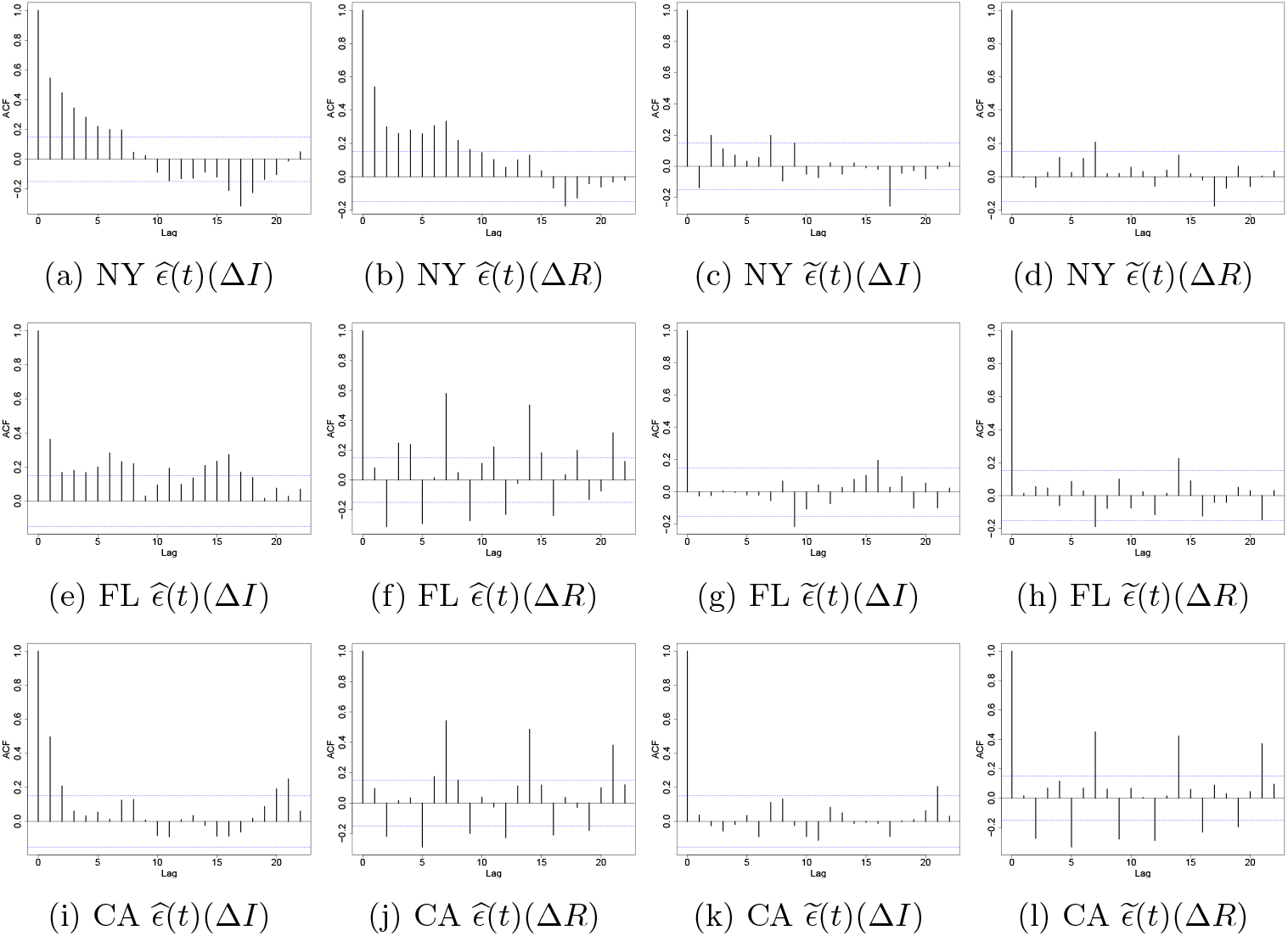
Auto-correlation plot of residuals by model 2.3 (left) and model 3 (right)

For a stationary time series *X_t_*, the auto-covariance function (ACVF) at lag *h* is defined as

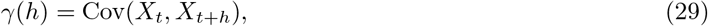

and the auto-correlation function (ACF) at lag *h* is defined as

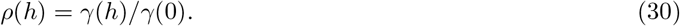

Let 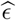 and 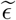 be the residuals from models 2 and 3, respectively. We use ACF to measure the degree of dependence among the residual processes at different times. Figure 8 shows the ACF of these residual time series. As can be seen from these plots, in all states, there is a statistically significant auto-correlation in the process 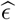 at different times while this temporal dependence has been reduced significantly in the residuals of model 3 which confirms the applicability of VAR modeling for the considered data sets.

## Appendix C: Additional Results for U.S. Counties

**Fig 9:**
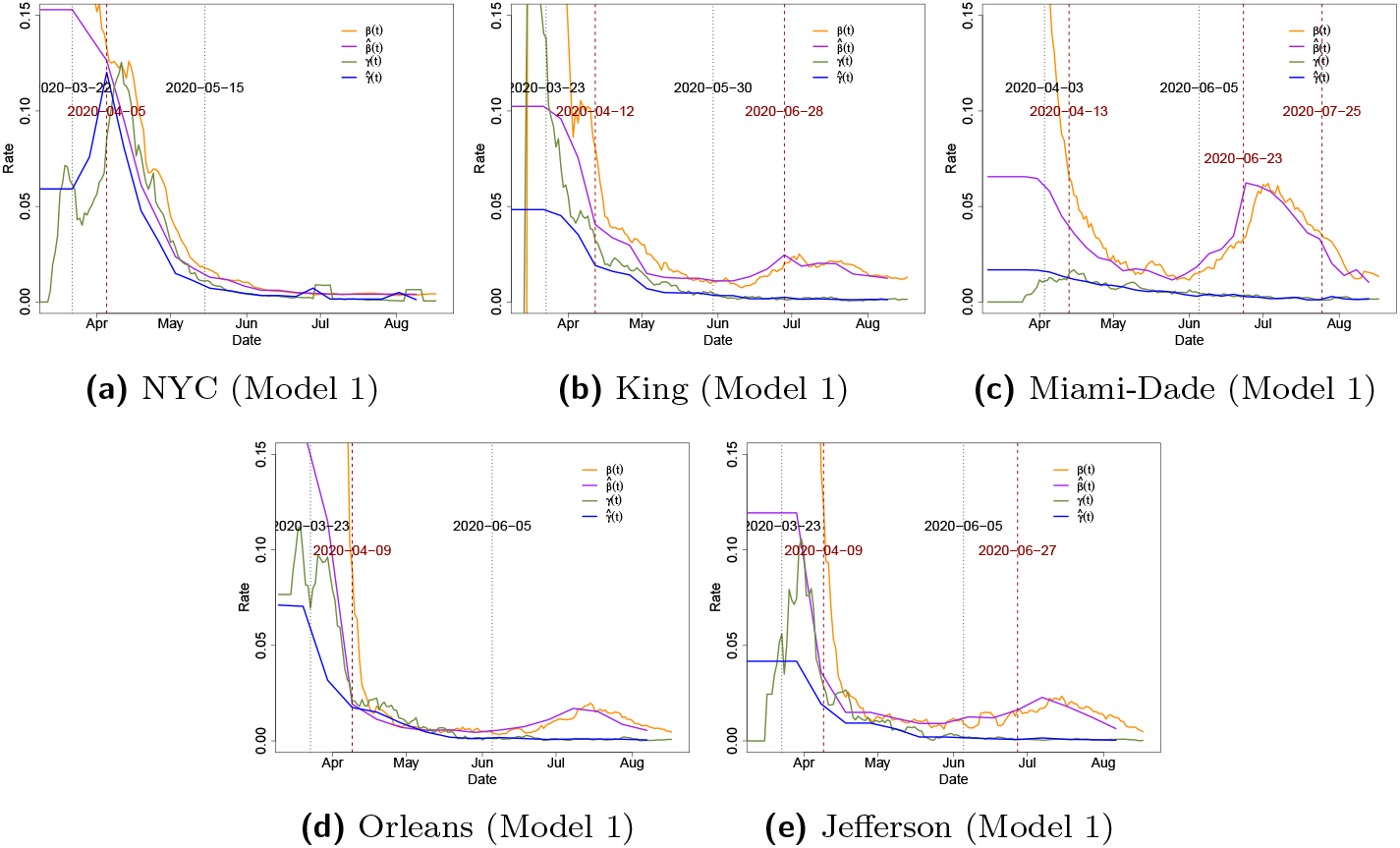
The 7-day moving average of observed (orange and green) and estimated (purple and blue) transmission rate and recovery rate in counties/cities/parishes. The vertical black dotted line indicates the statewide “stayat-home” order begin date or reopening begin date in the corresponding state. The vertical dark red dashed line indicates the estimated change time point in the county/city/parish.

**Fig 10:**
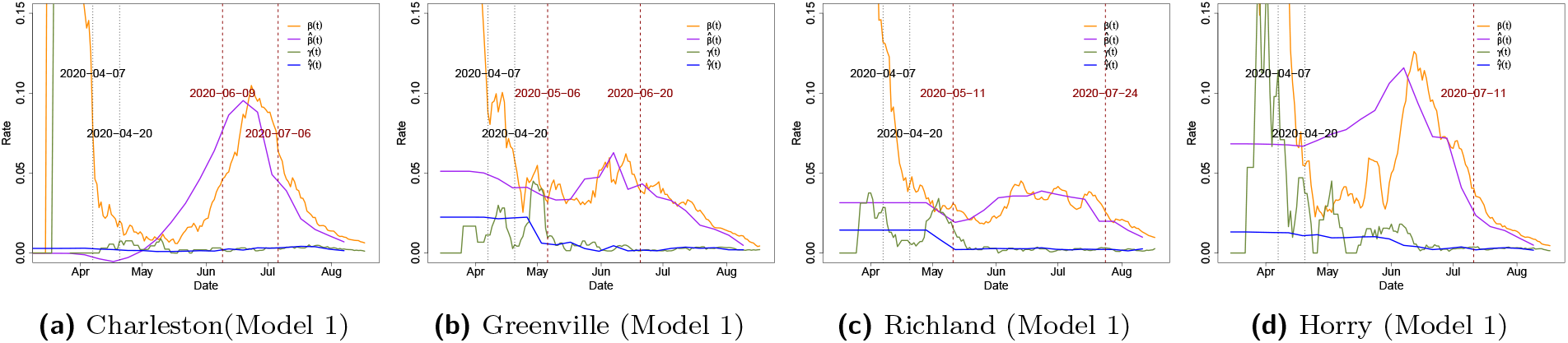
The 7-day moving average of observed (orange and green) and estimated (purple and blue) transmission rate and recovery rate in four counties in South Carolina. The vertical black dotted line indicates the statewide “stay-at-home” order begin date or reopening begin date in the corresponding state. The vertical dark red dashed line indicates the estimated change time point.

Few additional plots and tables related to the results of applying our method to some U.S. counties are provided in this section. The 7-day moving average of transmission *β*(*t*) and recovery *γ*(*t*) rates are provided in Figure 9 and 10. Further, the results in Table 16 indicate that the neighboring counties are making an significant impact on estimating and forecasting the transmission dynamic. Finally, Figure 11 shows the ACF of these time series in three counties: Orleans, Charleston and Richland.

**Table 16.** Spatial effect estimation in model 2.

|  | Model | Estimate | P-value | Confidence interval |
| --- | --- | --- | --- | --- |
| Orleans | Model 2.1 | 0.2651 | 0.00142 | ( 0.1032 , 0.4271 ) |
|  | Model 2.2 | 0.273 | 0.00135 | ( 0.1069 , 0.4391 ) |
|  | Model 2.3 | 0.2835 | 0.00138 | ( 0.1107 , 0.4562 ) |
|  | Model 2.4 | 0.3236 | 0.00050 | ( 0.1427 , 0.5046 ) |
| Charleston | Model 2.1 | 0.5843 | 2.64e-08 | ( 0.3832 , 0.7854 ) |
|  | Model 2.2 | 0.693 | 7.55e-11 | ( 0.4912 , 0.8949 ) |
|  | Model 2.3 | 0.9494 | 6.60e-21 | ( 0.765 , 1.1338 ) |
|  | Model 2.4 | 0.7831 | 5.18e-07 | ( 0.4828 , 1.0833 ) |
| Richland | Model 2.1 | 0.361 | 4.92e-13 | ( 0.2675 , 0.4546 ) |
|  | Model 2.2 | 0.3666 | 2.66e-13 | ( 0.2727 , 0.4604 ) |
|  | Model 2.3 | 0.7234 | 3.28e-24 | ( 0.5963 , 0.8506 ) |
|  | Model 2.4 | 0.6278 | 3.72e-15 | ( 0.4796 , 0.776 ) |

**Fig 11:**
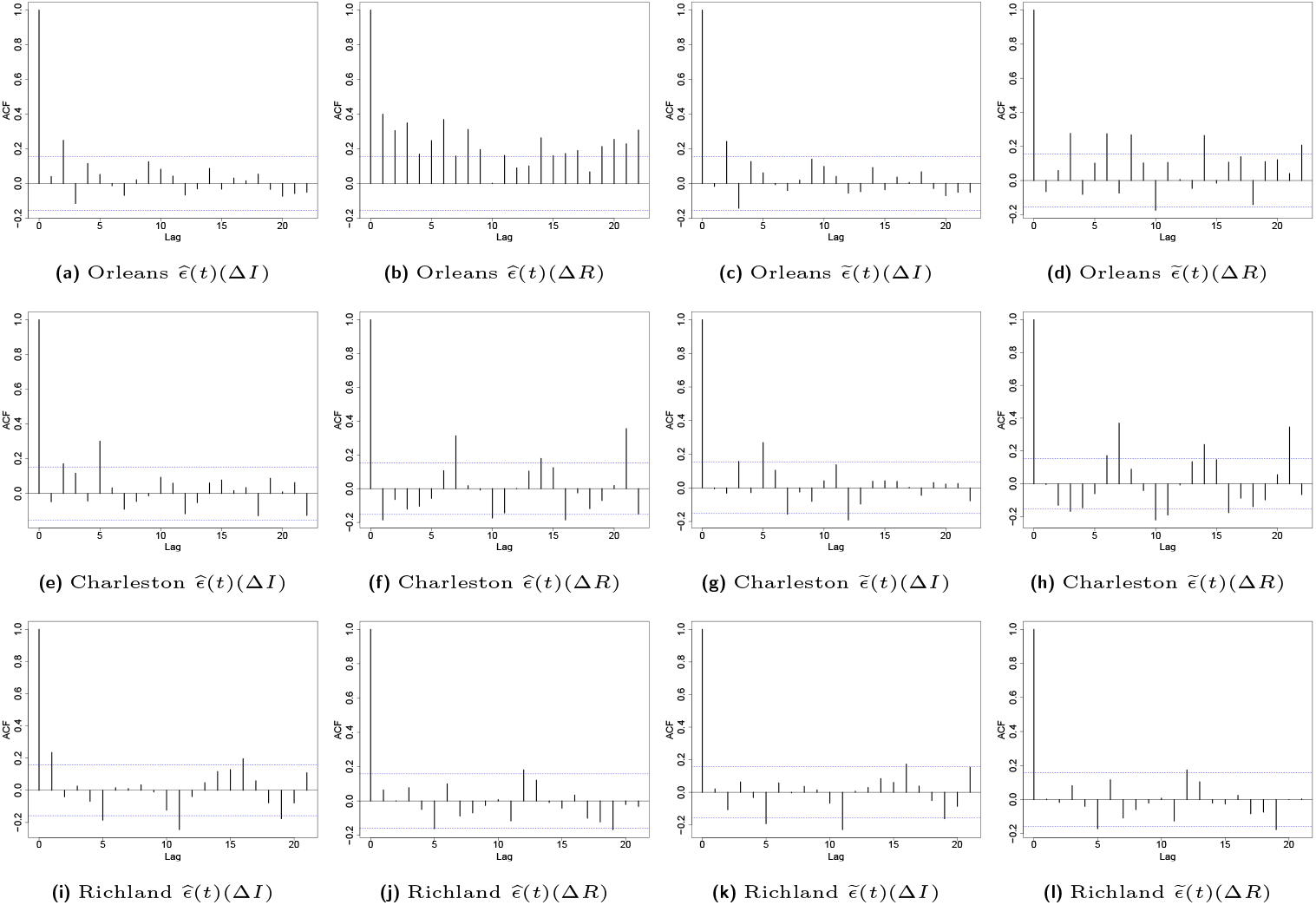
Auto-correlation plot of residuals by model 2.3 (left) and model 3 (right). County level.

## Footnotes

1 https://github.com/ybai69/COVID-19-Change-Point-Detection

2 https://www.cdc.gov/coronavirus/2019-ncov/cases-updates/commercial-lab-surveys.html

